# Frontoparietal Hyperconnectivity as a Single-Subject Neural Signature of Chronic Tinnitus

**DOI:** 10.64898/2026.11.20.26368561

**Authors:** Payam S. Shabestari, Shagun Ajmera, Elva Arulchelvan, Feifan Chen, Milena Engelke, Jessica Fritzsche, Namitha Jain, Emilia Kaniewska, Tobias Kleinjung, Hongyu Lu, Lauren Ralston Ruesch, Elinor C. Riegger, Nathan Shields, Susanne Staudinger, Beat Tödtli, Stephan M. Wolpert, Nina Wunder, Anusha Yasoda-Mohan, Fatima T. Husain, Julia Campbell, Dirk De Ridder, Berthold Langguth, Winfried Schlee, Sven Vanneste, Christopher R. Cederroth, Patrick Neff

## Abstract

Chronic tinnitus, the persistent perception of phantom sound, affects 14 to 15% of adults yet lacks a validated neurophysiological biomarker, in part because group-average EEG analyses obscure the individual heterogeneity that defines the condition. In a multi-site case-control study of 544 participants (268 with tinnitus, mean age 45.4 ± 16.1 years, 52% female) across seven independent international sites, we applied normative modelling to quantify how far each person’s resting-state EEG deviates from age- and hearing-adjusted healthy norms. Fusing spectral-power and connectivity deviations, a classifier detected tinnitus with an area under the curve of 0.832 under site-grouped cross-validation of precomputed normative features. The single most discriminative feature, rank-stable across all seven sites, was theta-band right orbitofrontal (*pars orbitalis*) connectivity, which dominated attribution by roughly an order of magnitude. Theta hyperconnectivity spanned frontoparietal and dorsal attention networks while primary auditory cortex showed negligible involvement, favouring failed top-down gating over bottom-up auditory overactivity. Features identifying tinnitus presence were orthogonal to symptom severity. These findings reframe chronic tinnitus as a frontoparietal attention-network disorder and nominate right orbitofrontal theta hyperconnectivity as a site-robust diagnostic feature. Data were collected within the TIDE project (ClinicalTrials.gov NCT06520865).

## I. Introduction

Tinnitus, the persistent conscious perception of sound in the absence of an external acoustic source, affects approximately 14 to 15% of adults worldwide, with around 2% experiencing symptoms severe enough to impair daily functioning [1]. It is among the most prevalent chronic sensory complaints and carries elevated rates of depression, anxiety, sleep disturbance, and cognitive decline [2]. Following international consensus [3], “tinnitus” (the percept alone) is distinguished from “tinnitus disorder” (the associated distress or functional disability). No symptom-modifying treatment exists, and available approaches target secondary distress rather than the underlying neural disturbance [4], [5]. A central reason for this stagnation is the absence of a validated, objective neural biomarker: without a reliable brain-based marker of the phantom sound percept, patient stratification, target identification, and treatment monitoring remain tied to subjective measures [6], [1], [7].

Chronic tinnitus is increasingly understood as an emergent property of large-scale network reorganisation rather than a localised auditory-cortex pathology [8], [6]. Peripheral deafferentation triggers maladaptive plasticity, from altered thalamocortical synchrony [9] to aberrant top-down gating by frontal circuits that normally suppress persistent, low-salience signals [10], [11], and resting-state studies report hyperconnectivity among the auditory, default-mode, and attention networks [12], [8], [13]. These accounts converge on a distributed, top-down disturbance rather than a single locus, but the bulk of previous data in humans has not been robustly replicated, and no single approach exists to detect tinnitus at the individual level.

Supervised classifiers applied to band power, source-space oscillatory features, and deep contrastive representations can distinguish the tinnitus brain state from controls with high reported accuracy: a linear support-vector machine separated 129 patients from 142 controls at 94% test accuracy [14], and a multi-band contrastive deep network achieved subject-independent diagnosis across 187 patients and 80 controls [15]. These estimates derive almost exclusively from single-site, within-cohort validation, however, where shared hardware, participant pool, and noise covariance inflate apparent performance beyond what transfers to new sites [16]. Two structural limitations therefore prevent existing classifiers from serving as clinical biomarkers. The first is cross-site generalisation: differences in amplifier hardware, montage, room acoustics, and operator protocol introduce batch effects that can rival the tinnitus signal itself [17], and although harmonisation methods such as *ComBat* partially correct them, including for resting-state EEG features specifically [18], no EEG tinnitus classifier has been validated under held-out site conditions. The second is neurobiological heterogeneity: patients vary widely in tinnitus laterality, chronicity, associated hearing loss, and psychological burden. These dimensions are weakly correlated and likely reflect distinct pathophysiology [19], [1], yet conventional case-control analyses estimate a single average difference and discard the individual variation needed for subtyping [20]. Normative modelling offers a principled alternative. It characterises a healthy reference distribution to yield per-subject deviation scores relative to age- and hearing-corrected norms [20], [21]. Applied to resting-state EEG, it has mapped individual source-connectivity deviations in Parkinson’s disease that scale with clinical severity [22], localised epileptogenic tissue in focal epilepsy [23], [24], and, most directly analogous here, predicted antidepressant response from individual EEG-connectivity deviations [25]. It has not yet been extended to tinnitus.

Here we address both limitations by applying normative modelling to resting-state EEG in a multi-site case-control design of 544 participants (268 chronic tinnitus, 276 controls) across seven independent international sites in the TIDE consortium [26]. The TIDE protocol additionally acquired task paradigms, including auditory oddball and gap pre-pulse inhibition of acoustic startle (GPIAS) recordings, the present work focuses on the resting-state EEG, with the task paradigms analysed in companion papers. Individual deviation scores, corrected for age, sex, and hearing thresholds, were estimated from controls alone under leave-one-site-out (LOSO) validation, and rather than committing to a fixed pipeline we benchmarked feature representations, connectivity measures, and classifiers within the same scheme, projecting TreeSHAP [27] attribution onto the cortical surface to localise the dominant signature. We ask three questions: (i) can normative EEG deviations identify chronic tinnitus at the single-subject level across sites unseen during training? (ii) which circuits and frequency bands carry the diagnostic signal? and (iii) do the most discriminative features converge on a stable, cross-site neurophysiological signature?

## II. Results

### A. PLI normative models are well-calibrated and reveal a hyperconnectivity asymmetry

Bayesian linear regression normative models [28], fitted to controls under leave-one-site-out (LOSO) cross-validation, were well calibrated for the two phase-lag-index (PLI) connectivity modalities: regional and global PLI Z-scores had median feature means of 0.043 and 0.044 (median SDs 1.025 and 1.031), within rounding of the *N* (0, 1) targets, with 5.4% of values exceeding |1.96| (nominal 5.0%), indicating near-nominal tails and negligible mean bias (Fig. 1a). Spectral power instead showed a systematic positive shift (median feature mean 0.711, 6.1% exceeding |1.96|), reflecting a LOSO-fold extrapolation bias when a site is absent from training, because power in isolation achieved only AUC = 0.583 under the same scheme, the inflated Z-scores carry no discriminative signal and all primary conclusions are drawn from the well-calibrated PLI modalities.

**Fig. 1.**
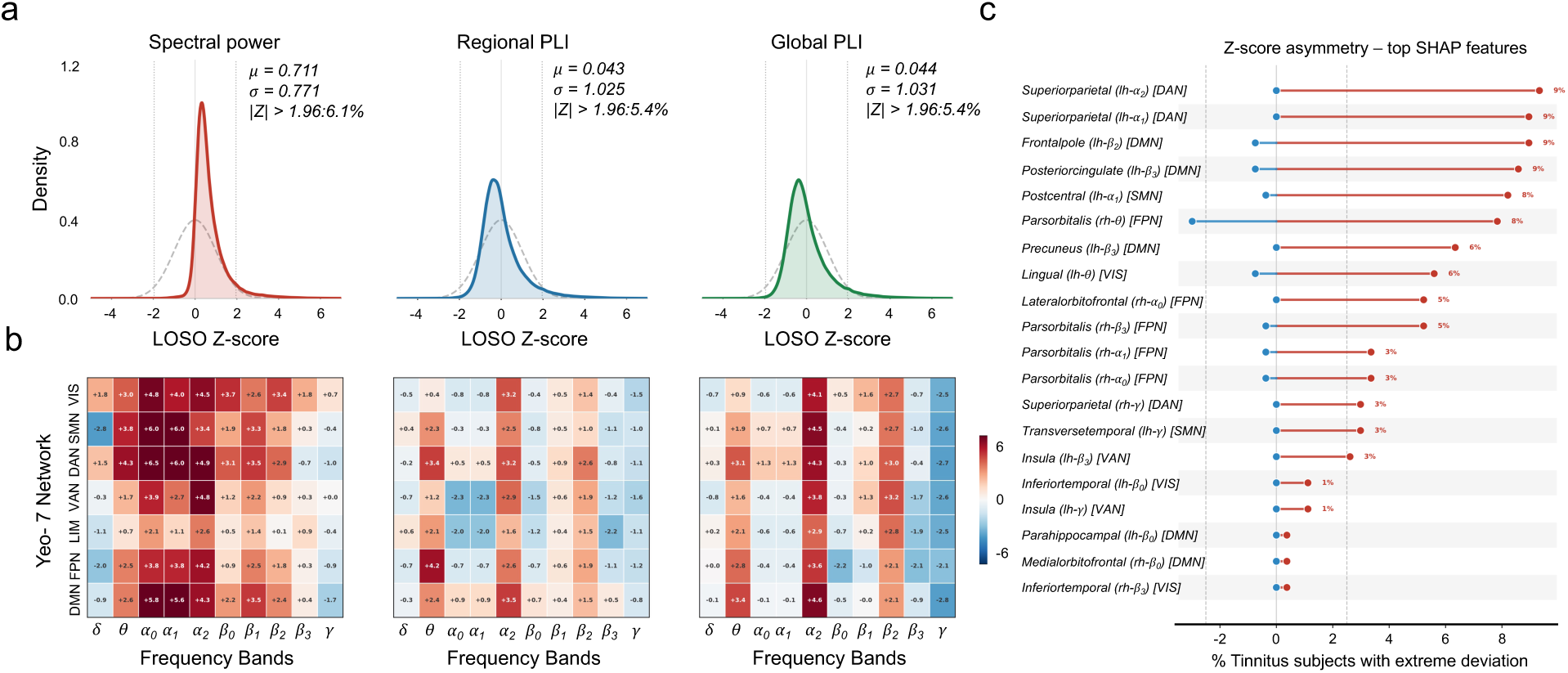
Normative model calibration, tinnitus deviation distributions, and individual-level prevalence of extreme deviations. (**a**) Per-modality calibration of control LOSO Z-scores (KDE of pooled within-feature Z-scores, dashed: *N* (0, 1) reference, dotted verticals: ±1.96). Regional PLI (median feature mean = +0.043, median SD = 1.025, 5.4% exceeding |1.96|) and global PLI (median mean = +0.044, median SD = 1.031, 5.4%) closely follow *N* (0, 1), spectral power shows a systematic positive shift (median mean = +0.711, median SD = 0.771, 6.1%). (**b**) Network × frequency-band prevalence difference maps for spectral power, regional PLI, and global PLI. Each cell shows the difference in the percentage of subjects with hyperdeviations (*Z >* +1.96) between tinnitus patients and controls (tinnitus − controls, in percentage points). Positive values (red) indicate cells where a disproportionate fraction of tinnitus subjects exceed the normative threshold relative to controls, negative values (blue) indicate the reverse. Under a well-calibrated normative model, controls are expected to exceed the threshold in approximately 5% of subjects, so a positive difference reflects a genuine tinnitus-specific deviation pattern. The black border highlights the FPN × *θ* cell, which shows the largest consistent excess across modalities. (**c**) PLI node-strength Z-score asymmetry: proportions of tinnitus subjects exceeding +1.96 (hyperdeviation) vs. −1.96 (hypodeviation) per feature, showing a large hyperconnectivity ratio.

Network-band prevalence maps (hyperdeviation excess, tinnitus minus controls, per Yeo-7 network [29] × band, Fig. 1b) showed two features: a spatially focal theta excess in the frontoparietal control network (FPN, 7.8% of tinnitus vs. 3.5% of controls in regional PLI, Δ = 4.2 percentage points, the largest theta effect of any network), and a broad, non-specific upper-alpha (*α*_2_) elevation appearing across most networks in both power and connectivity. This juxtaposition motivates the multivariate analysis: threshold counts are sensitive to any diffuse shift, and only continuous Mahalanobis deviations resolve the specific axis. When those are tested with FDR correction (Fig. 3a), four cells survive, three in theta (FPN, DAN, DMN) plus VIS×*β*_2_, with FPN×*θ* dominant by two orders of magnitude, whereas the visually prominent *α*_2_ elevation reaches significance in no network (FPN×*α*_2_: Δ = 0.370, *p*_FDR_ = 0.198). Tinnitus deviations were also strongly directional: 5.3% of PLI Z-scores exceeded +1.96 but only 0.3% fell below −1.96 (Fig. 1c), an asymmetry most pronounced for the top SHAP [30] feature (*parsorbitalis*-rh × theta), whose node-strength Z-score had a narrow control distribution (SD = 0.42) but a broad rightward-shifted tinnitus distribution (SD = 1.38), indicating individual heterogeneity in orbitofrontal connectivity absent in the healthy population.

### B. Band-resolved multi-modal PLI fusion outperforms single-modality and flat-feature alternatives

Because no single feature representation, connectivity measure, or classifier is guaranteed to generalise across sites, we treated the pipeline as a search space, benchmarking a systematic grid of scenarios under the same 7-site LOSO protocol and reporting the best configuration plus an ensemble over the strongest scenarios. The primary scenario, a LightGBM classifier [31] on the 680 band-resolved multi-modal Mahalanobis deviation features (PLI, pre-processing Level 2), achieved AUC = 0.819, balanced accuracy 74.7%, sensitivity 68.5%, and specificity 80.9% (Fig. 2d), a soft-voting ensemble over the top-5 scenarios raised AUC to 0.832 (Fig. 2c). Ablation across four axes isolated the drivers (Fig. 2a-b): the band-resolved 680-dimensional representation (AUC = 0.830) far outperformed the band-averaged 68-dimensional version (0.579), raw deviation connectivity (0.690), and residual connectivity (0.517). PLI (0.819) surpassed PLV (0.737) and coherence (0.695) [32], confirming the value of volume-conduction-robust synchrony. LightGBM (0.830) outperformed random forest (0.735) and SVM (0.707), and all single-modality baselines were inferior to the three-modality fusion (graph PLI 0.793, regional PLI 0.672, global PLI 0.624, aperiodic slope/offset 0.596, power 0.583). Hyperparameter optimisation via `Optuna` [33] did not improve over fixed parameters, indicating the default LGBM configuration was already near-optimal.

**Fig. 2.**
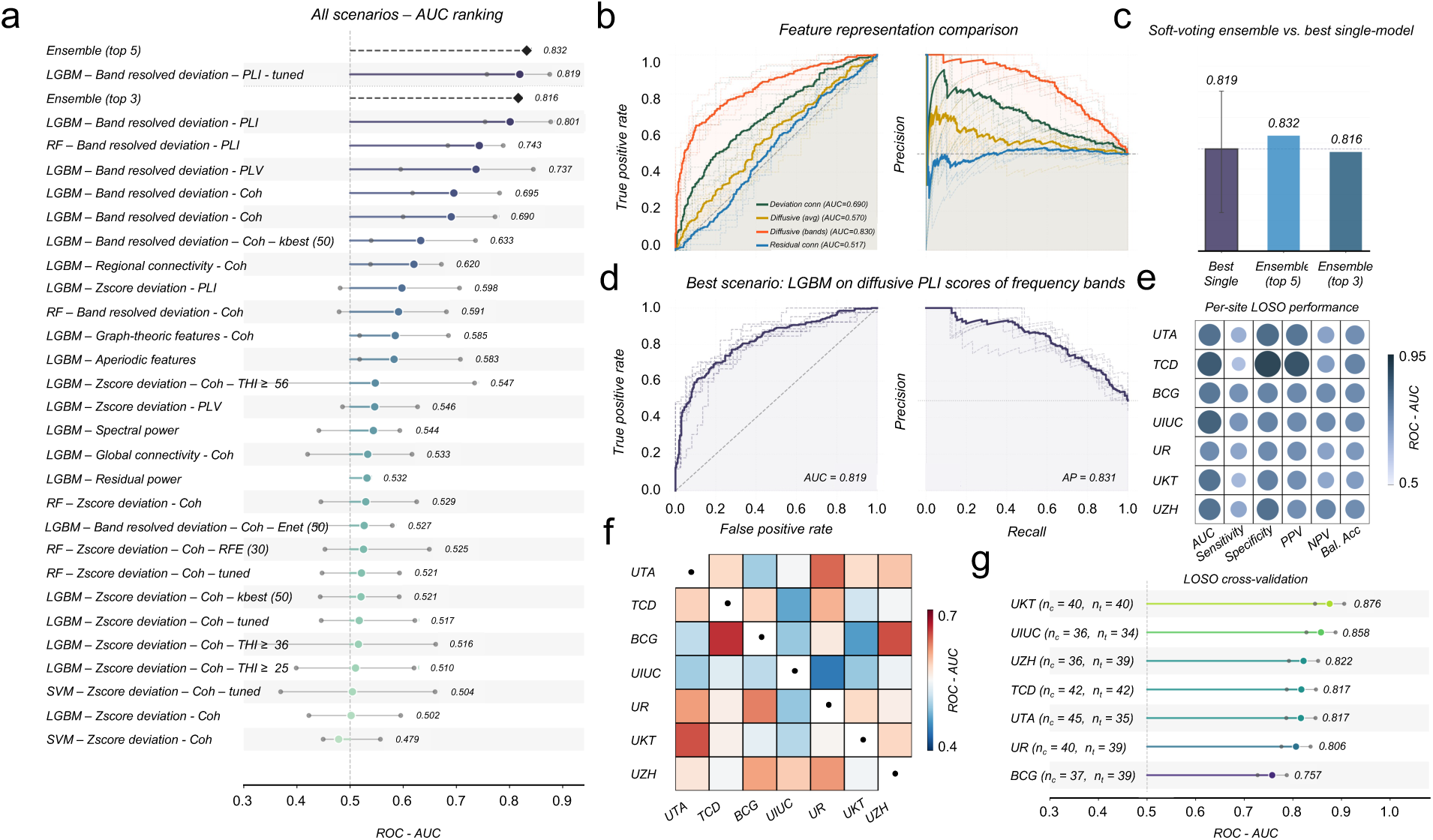
Classification performance: ablation, site generalisation, and cross-site transfer. (**a**) AUC across all ablation axes: feature representation (band-averaged 68-dim vs. band-resolved 680-dim), connectivity measure (PLI, PLV, COH), classifier (LGBM, RF, SVM), and single-modality baselines. Each bar aggregate LOSO AUC over site folds. (**b**) Feature representation comparison: residual connectivity, deviation connectivity, band-averaged Mahalanobis, and band-resolved Mahalanobis. (**c**) Soft-voting ensemble AUC (top-3 and top-5 scenarios) vs. best single model. (**d**) ROC and precision-recall curves for the best scenario (band-resolved PLI LGBM): aggregate (solid), per-site LOSO folds (dashed). (**e**) Per-site classification performance across five metrics — sensitivity (tinnitus recall), specificity (control recall), AUC, positive predictive value (PPV), and negative predictive value (NPV), and balanced accuracy, showing consistently high specificity and AUC across all sites, with greater variability in sensitivity and predictive values reflecting site-level differences in class balance and subject heterogeneity. (**f**) 7×7 cross-site transfer matrix (AUC) when training on a single site and testing on each other site, near-chance values (0.45–0.66) contrast with LOSO AUC = 0.819, establishing that multi-site aggregation is essential. (**g**) Per-site AUC from the primary scenario (LGBM, band-resolved PLI, 680 features), with bootstrap 95% CIs (1000 iterations). All seven sites significantly exceed chance.

### C. Multi-site LOSO yields strong, consistent classification across all seven held-out sites

Under the primary scenario, per-site LOSO AUC ranged from 0.757 (BCG) to 0.876 UKT), with all seven sites individually exceeding 0.750 and bootstrap 95% confidence intervals confirming every site exceeded chance (Table I, Fig. 2e,g). This site-level range is substantially narrower than in comparable single-site EEG tinnitus studies, demonstrating that the normative pipeline learns features generalisable across hardware, noise covariance, and other situational factors. Individual-level error analysis showed that false negatives (*n* = 85) and false positives (*n* = 52) span the full probability range rather than clustering at the decision boundary, with per-site rates varying and UKT showing the largest asymmetry (Supplementary Fig. S2).

**TABLE I.** Per-site LOSO classification performance. (primary scenario: LGBM, band-resolved PLI, 680 features). PPV = positive predictive value, NPV = negative predictive value.

| **Site** | *n*<sub>ctrl</sub> / *n*<sub>tin</sub> | **AUC** | **Sens.** | **Spec.** | **PPV** | **NPV** |
|---|---|---|---|---|---|---|
| UTA | 45 / 35 | 0.817 | 0.686 | 0.822 | 0.750 | 0.771 |
| UIUC | 36 / 34 | 0.858 | 0.735 | 0.778 | 0.758 | 0.757 |
| TCD | 42 / 42 | 0.817 | 0.643 | 0.786 | 0.750 | 0.688 |
| BCG | 37 / 39 | 0.757 | 0.692 | 0.757 | 0.750 | 0.700 |
| UZH | 36 / 39 | 0.822 | 0.667 | 0.833 | 0.813 | 0.698 |
| UKT | 40 / 40 | 0.876 | 0.625 | 0.925 | 0.893 | 0.712 |
| UR | 40 / 39 | 0.806 | 0.744 | 0.775 | 0.763 | 0.756 |
| Mean |  | 0.822 | 0.685 | 0.811 | 0.782 | 0.726 |

To probe whether single-site training can transfer to unseen sites, we constructed a 7 × 7 cross-site transfer matrix (Fig. 2f), using a random forest rather than LightGBM because each single-site set contains only 35 to 90 subjects, far below the size for which LGBM was tuned, if anything this yields a conservative estimate of single-site failure, since LGBM would overfit more severely. Cross-site AUC ranged from 0.45 to 0.66, far below the multi-site LOSO values, establishing that multi-site aggregation is essential rather than merely convenient: the signal-to-noise ratio of any individual site is insufficient to train a generalisable classifier.

### D. The frontoparietal network shows uniquely selective theta-band hyperdeviation

Across the 7 × 10 network×band cells, Mann-Whitney tests with FDR correction revealed a spatially and spectrally specific deviation pattern (Fig. 3a). The dominant effect was FPN×*θ* (*p*_FDR_ < 0.0001, Δ = 0.404), with secondary effects at DAN×*θ* (*p*_FDR_ < 0.001, Δ = 0.429), DMN×*θ* (*p*_FDR_ < 0.05, Δ = 0.301), and VIS×*β*_2_ (*p*_FDR_ < 0.05, Δ = 0.272), all elevated in tinnitus. SMN, VAN, and LIM showed no significant deviation in any band, and, of special interest, primary auditory ROIs (*transversetemporal*, *superiortemporal*) showed no group difference, indicating that tinnitus is not characterised by auditory-cortex hyperdeviation once hearing loss is controlled for. Theta dominance was not uniform across frontal networks: FPN showed a *p*_FDR_ nearly 100× smaller than DAN, pointing to preferential involvement of prefrontal and parietal association cortex, with a weak, non-significant extension into *α*_2_ (*p*_FDR_ = 0.198, Δ = 0.370).

**Fig. 3.**
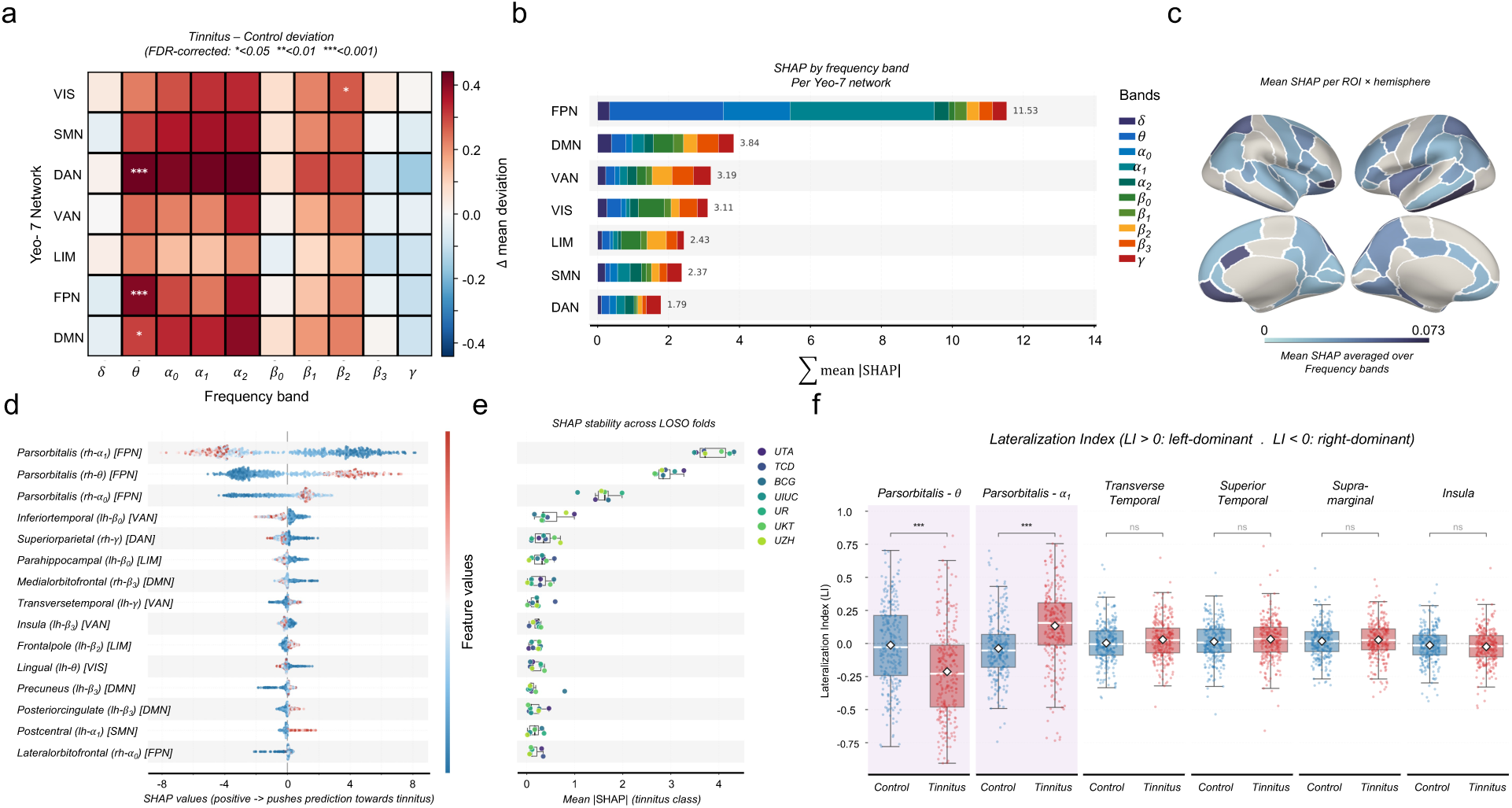
Network-level deviation signature, TreeSHAP feature attribution, and hemispheric lateralisation. (**a**) Network × frequency-band deviation heatmap: mean diffusive Mahalanobis deviation difference (tinnitus − controls) per Yeo-7 network [29] and frequency band. Colour intensity: effect size. Stars: FDR-corrected Mann-Whitney *U* significance (*^∗^p*_FDR_ < 0.05, *^∗∗^p*_FDR_ < 0.01, *^∗∗∗^p*_FDR_ < 0.001). FPN × *θ* is the dominant effect by two orders of magnitude. (**b**) Network-level SHAP importance (mean *|* SHAP| summed over member ROIs and bands). FPN contributes the majority, DAN is second. (**c**) Cortical surface map of mean |SHAP| per ROI averaged across frequency bands (analogous to an fMRI whole-brain contrast map). Bilateral prefrontal gradient, maximum at right *parsorbitalis* (see Supplementary Fig. S6 for band resolved surface maps). (**d**) Mean |SHAP| for the top 12 features (tinnitus class), aggregated across all seven LOSO folds. *Parsorbitalis*-rh dominates by a factor of *>* 1.3× over rank 2. (**e**) Per-fold SHAP stability for the top 15 features (box: fold distribution, dots: individual held-out sites). *Parsorbitalis*-rh × *θ* and × *α*_1_ are rank-stable across all 7 folds. (**f**) Lateralisation index (LI) distributions for *parsorbitalis* in theta and *α*_1_ bands (tinnitus vs. controls). In theta, tinnitus subjects show accentuated right dominance (LI_tin_ = −0.213 vs. −0.012, *p*_FDR_ = 8.3 × 10^−12^), in alpha-1, the direction reverses to left dominance (LI_tin_ = +0.134 vs. −0.037, *p*_FDR_ = 1.6 × 10^−16^). LI *>* 0: left-hemisphere dominant, LI < 0: right-hemisphere dominant. LI distributions for auditory and auditory-adjacent ROIs (*transversetemporal*, *superiortemporal*, *supramarginal*, *insula*). No significant group difference survives FDR correction (all *p*_FDR_ *>* 0.21), confirming hemispheric asymmetry is specific to the prefrontal signature.

### E. Right pars orbitalis anchors classification with perfect stability across held-out sites

TreeSHAP [27] under LOSO identified *parsorbitalis-rh* as the top-ranked feature in every fold (Fig. 3d-e). At band resolution the three highest-attribution features were *parsorbitalis*-rh ×*α*_1_ (mean |SHAP| = 3.86), ×*θ* (2.89), and ×*α*_0_ (1.59). The drop to rank 4 (*inferiortemporal*-lh ×*β*_0_: 0.49) was sharp, with rank 1 exceeding rank 4 by 7.9× and the combined top-three *parsorbitalis*rh features by 17×, accounting for *≈* 52% of cumulative attribution. At the network level, FPN contributed the largest share, followed by DAN and a smaller DMN contribution (Fig. 3b), the surface projection shows a right-lateralised orbitofrontal hotspot, with secondary attribution distributed across medial-temporal, parahippocampal, inferior-temporal, parietal, and cingulate regions, and lowest attribution in occipital and primary sensory areas (Fig. 3c).

### F. Pars orbitalis shows frequency-selective hemispheric reversal

We asked whether the orbitofrontal signature carries a hemispheric bias, and whether any asymmetry is specific to it, computing a per-subject lateralisation index (LI, Methods) for *parsorbitalis* and, as a comparison set, four ROIs spanning the auditory hierarchy and salience circuitry (Fig. 3f). *Parsorbitalis* showed frequency-selective reversal: in theta both groups were right-dominant but tinnitus more strongly so (LI_tin_ = −0.213 vs. −0.012, *p*_FDR_ < 0.0001), while in *α*_1_ the pattern reversed to left-dominance in tinnitus (LI_tin_ = 0.134 vs. −0.037, *p*_FDR_ < 0.0001), indicating distinct oscillatory mechanisms that cannot be reduced to a single hemisphere or band. None of the four auditory and salience ROIs (*transversetemporal*, *superiortemporal*, *supramarginal*, *insula*) showed a significant lateralisation difference after FDR correction, reinforcing that the dominant reorganisation is prefrontal and attention-network-based rather than auditory or salience-lateralised.

### G. EEG oscillatory deviations detect tinnitus presence but not symptom burden

Despite accurate detection (AUC = 0.819), regression of THI and TFI from the same 680 features yielded strongly negative *R*^2^ across every model and target: under site-stratified cross-validation Ridge reached *R*^2^ = −0.60 (THI) and −0.45 (TFI), and LightGBM did no better (−0.67, −0.54), all below zero and therefore worse than a constant group-mean baseline. Fold values were highly variable (THI range −1.26 to −0.14), indicating systematic rather than stochastic failure (Supplementary Fig. S3). The strongest single-feature association reached only *ρ* = +0.15 (THI) and +0.10 (TFI), with none surviving FDR correction, confirming that no single neural metric tracks distress severity.

### H. Normative deviations are sparse, topographically selective, and individually heterogeneous

Feature-level prevalence maps (proportion with *|Z| >* 1.96 per ROI-band, aggregated by network, Fig. 1b) showed a sparse, selective hyperdeviation pattern: most features sat near the *≈* 5% chance rate, with a subset reaching 10 to 16%, localised in regional PLI to FPN ROIs in theta and alpha (*parsorbitalis-lh α*_2_: 12.7%, *θ*: 10.8%), paralleling the SHAP-ranked features. Hypodeviation was near-zero (0.3% vs. 2.5% expected) while hyperdeviation reached 5.3%, so tinnitus deviations are exclusively upward at every scale. Yet 96 to 98% of tinnitus subjects showed at least one extreme deviation in regional PLI, the hallmark of a heterogeneous condition: nearly every subject carries a neural fingerprint of abnormality, but in a subject-specific combination of ROIs and bands, so it is the spatial-spectral identity of the deviating features, captured by the trimodal Mahalanobis representation, not their raw count, that provides diagnostic value. Age analysis showed no significant age-deviation correlation in tinnitus (global mean *r_s_*= 0.053, *p* = 0.389) and no age×group interaction surviving FDR correction, indicating an age-stable signature, notably *parsorbitalis*-rh ×*θ* showed a modest positive age trend in controls (*r_s_* = 0.225, *p*_FDR_ = 0.003) absent in tinnitus (*r_s_* = 0.134, *p*_FDR_ = 0.185), suggesting the normal age-related rise in orbitofrontal theta connectivity is obscured by the tinnitus-related elevation (Supplementary Fig. S5a). Trimodal scatterplots confirmed power and PLI Z-scores are imperfectly correlated (*|r| ≈* 0.3 to 0.5), validating the Mahalanobis fusion (Supplementary Fig. S5b).

### I. Preprocessing without ICA maximises classification performance

Across three preprocessing levels under 7-site LOSO, Level 2 (*autoreject* [34] epoch rejection and interpolation, without ICA) achieved the highest AUC (0.832), outperforming the more lenient Level 1 (no artefact rejection, 0.646) and the more aggressive Level 3 (*autoreject* with *ICLabel* ICA removal, 0.741, Supplementary Fig. S3b). Omitting rejection cost 0.186 AUC while adding ICA removal still cost 0.091, placing the optimum at intermediate stringency. This non-monotonic pattern replicates Bomatter et al. [35] and validates Level 2: standard epoch cleaning suffices, and aggressive ICA removal discards signal rather than noise.

## III. Discussion

We report four principal findings. First, a normative, multi-modal fusion model detects chronic tinnitus from resting-state EEG at the single-subject level and generalises to entirely held-out recording sites, moving classification into a clinically informative range. Second, the discriminative signal converges on a single anatomical anchor, theta-band connectivity of the right pars orbitalis, embedded in the frontoparietal control network (FPN), while auditory cortex shows no deviation once hearing loss is accounted for. Third, the network deviation that detects the percept is statistically orthogonal to subjective distress, indicating that detection and burden are separable neural constructs. Fourth, two methodological factors, artefact-rejection stringency and multi-site training, are decisive for whether a biomarker is recoverable at all.

### A. A normative multi-modal fusion framework reaches the clinical-utility range

The three-modality Mahalanobis normative model achieved AUC = 0.819 (ensemble: 0.832) across 544 participants at seven independent international sites under LOSO cross-validation, with per-site AUC ranging from 0.757 to 0.876. To our knowledge all prior EEG tinnitus classifiers were single-site and none reported held-out multi-site validation [15], [14], so a direct AUC comparison is inappropriate: within-site cross-validation inflates apparent generalisability by exploiting site-specific covariance. These results therefore establish the first tinnitus classifier validated across independent sites and the first detection estimate not inflated by within-site optimism. Two design choices explain the advance. Normative modelling first converts each feature into a Z-score relative to a healthy reference, decoupling individual deviation from site, age, sex, and audiometric confounds, multi-modal fusion then stacks power, regional node strength, and global node strength into a covariance space whose Mahalanobis distance captures joint deviations no single modality encodes. Together, cross-site validity, a stable neural correlate, and a short inexpensive recording outline a low-barrier route toward objective diagnosis complementing the questionnaires on which stratification currently depends. The area under the curve, which integrates performance across all thresholds, rather than any single operating point is the appropriate summary of clinical value: a fixed threshold trades sensitivity against specificity, so that specificity near 0.80 yields sensitivity near 0.68 and misses roughly one in three tinnitus cases. The operating point should therefore be matched to the intended use, favouring sensitivity for screening and specificity for confirmation, with Youden’s index (*J* = sensitivity+specificity − 1) offering one principled, application-tunable choice.

### B. Theta frontoparietal hyperconnectivity favours disrupted predictive coding and frontal gating

The single largest group deviation is a theta-band connectivity increase within the FPN, with a parallel elevation in the dorsal attention network (DAN, *p*_FDR_ < 0.001). This is consistent with the frontostriatal gating model [10], [11], in which ventromedial prefrontal cortex and nucleus accumbens normally suppress tinnitus-related activity at the thalamic reticular nucleus, when this fails the percept persists. Our data cannot test the subcortical link directly, but the sustained cortical theta engagement is what that model predicts at the cortical level: prefrontal and parietal association cortices left in continuous engagement with an internally generated percept the gate has not resolved, imposing an ongoing attentional demand that the DAN elevation mirrors. This emphasis is supported by earlier but underweighted EEG evidence of abnormal prefrontal coupling in chronic tinnitus [8] and of oscillatory disturbance beyond auditory cortex [12], [36], [9]. When individual deviations are modelled rather than group means, this prefrontal involvement emerges as the single most discriminative feature of the tinnitus brain state, the intrinsic-state counterpart of the task-evoked prefrontal over-recruitment that scales with perceptual intrusiveness [37].

One point needs care in interpretation. We measure phase synchrony, which quantifies how tightly two regions’ oscillations are coupled, not the direction of information flow. Elevated synchrony therefore means prefrontal and parietal cortices are more tightly phase-coordinated in tinnitus, on its own it does not indicate whether top-down or bottom-up signalling drives the coupling, a distinction that matters when relating our findings to predictive-coding accounts framed in terms of directional prediction error. Because midfrontal theta is a robust, transdiagnostic index of cognitive control that scales with conflict and difficulty [38], frontoparietal theta hyperconnectivity is most parsimoniously read as sustained over-engagement of the control network rather than its failure: circuitry transiently recruited to resolve conflict appears tonically recruited in chronic tinnitus, allocating control resources to a signal that is never behaviourally resolved. That mechanism is not tinnitus-specific, and we do not claim otherwise, what is specific is the fingerprint, a right pars orbitalis theta anchor with an intact auditory cortex, that a normative model assigns reproducibly to the individual. Whether that fingerprint separates tinnitus from other control-network disorders is a question our single-disorder design cannot answer.

The predictive-coding framework specifies why the recruitment might persist: tinnitus arises when the auditory generative model assigns abnormally high precision to spontaneous activity, so that compensatory neural noise is misread as sensory evidence [39], and the precision-weighting that sustains this is itself a frontoparietal function. Direct EEG evidence that patients with minimal hearing loss show a top-down subtype driven by maladaptive predictive coding rather than peripheral drive [40] is consistent with our theta hyperconnectivity as the expression of this aberrant precision-weighting. A complementary, band-specific reading is that prefrontal-parahippocampal theta loops progressively embed the percept in long-term memory, with the hippocampus maintaining tinnitus as an auditory object held in memory rather than acting solely as a limbic emotion generator [41]. This aligns with the beta-band medial-temporal deviations we observe (Fig 3d) and offers a concrete mechanism for chronification that our cross-sectional data are consistent with but cannot yet prove.

### C. Right pars orbitalis is the central tinnitus anchor

The most striking explainability result is the perfect fold stability of *parsorbitalis-rh*, which ranked as the most informative feature in every LOSO fold. This robustness across seven heldout sites is unprecedented in the tinnitus EEG literature and indicates a genuine, generalisable signature rather than an artefact, earlier work missed it because single-site univariate case-control analyses do not retain the individual multivariate deviation structure in which this feature is strongest. The right *parsorbitalis* is an orbitofrontal subdivision of the limbic-prefrontal circuit governing salience evaluation, emotional tagging, and prediction error [42], with connectivity to amygdala, anterior cingulate, and auditory association cortex placing it at the intersection of emotional processing and auditory gating. This position is coherent with the triple-network model, in which tinnitus emerges when interplay between salience, executive-control, and default-mode systems is disturbed so that a self-generated percept is repeatedly flagged as salient and cannot be released from attention [19]. The lateralisation is frequency-selective: theta showed accentuated right-hemisphere dominance in tinnitus (LI_tin_ = −0.213 vs. LI_ctrl_ = −0.012), whereas *α*_1_ (8.5 to 10.5 Hz) reversed to left-dominance (LI_tin_ = 0.134 vs. LI_ctrl_ = −0.037, *p*_FDR_ < 0.001), suggesting right-prefrontal theta reflects salience and gating while the *α*_1_ left-shift may index compensatory left-attentional recruitment. The same orbitofrontal locus was independently identified as a connectivity hub in a parallel resting-state MEG study [43], indicating that the signature recurs across modalities.

### D. Tinnitus is biased towards top-down, not bottom-up cortical alterations

A theoretically important null result is the complete absence of significant deviation in primary auditory cortex ROIs and connectivity (*transversetemporal*, *superiortemporal*) after controlling for hearing loss [44]. Auditory-cortex hyperactivity has historically been treated as a hallmark of tinnitus, inferred from animal models of deafferentation and reported in human M/EEG studies relating auditory-cortex gamma activity to the percept [36], [45]. These rarely controlled for audiometric confounds with the precision of a per-feature normative model. This does not imply the auditory cortex is uninvolved in generating the percept, only that residual auditory-cortex deviation does not distinguish tinnitus patients from hearing-loss-matched controls. It fits a gating-failure account in which the cortex produces the initial phantom signal after deafferentation, but chronic conscious perception depends on whether the OFC/FPN system suppresses it, so that the distinguishing feature of non-habituated patients is frontoparietal hyperconnectivity rather than auditory-cortex overactivity. Neuromodulation evidence reinforces this: stimulation confined to auditory cortex yields small, inconsistent benefits, whereas prefrontal targets more reliably shift the percept and are among the more consistently effective [1]. The network whose modulation most affects chronic tinnitus is the same frontoparietal system that dominates our classifier.

### E. Neural deviation and subjective burden are orthogonal

The signature indexes the percept, not its burden. All regression models were systematically worse than a constant predictor (Ridge-THI: *R*^2^ = −0.60, Ridge-loudness: *R*^2^ = −2.51), a genuine dissociation rather than weak signal: features that classify tinnitus presence cannot predict how loud or distressing it is. This matches the anatomy of our deviations, confined to attentional and cognitive-control networks (FPN, DAN) with no limbic locus surviving FDR correction, whereas tinnitus distress localises to salience, limbic, and default-mode circuitry [46], [47] that THI and TFI capture but our features do not. Subjects who deviated most strongly were not the most distressed, consistent with the heterogeneity of person-specific deviations within a diagnosis [28], [48]. A severity biomarker therefore cannot be built by aggregating deviation magnitudes, but requires the circuit-level mechanisms that govern whether an abnormal oscillatory state is appraised as distressing. Detection and severity monitoring thus demand different pipelines and population stratification, as illustrated by a companion paper by Lu et al. using a cortical adaptation of gap pre-pulse inhibition of the acoustic startle (GPIAS), where impaired inhibition correlated with tinnitus distress [49].

Perceptual character is equally decoupled. Misclassified subjects span the full predicted-probability range rather than clustering at the 0.5 boundary (Supplementary Fig. S2), and SHAP places false-negative subjects within the healthy range of frontoparietal PLI deviation, so a subset of tinnitus subjects genuinely present as neural controls. These false negatives are a mechanistically distinct subgroup, not borderline errors: median predicted probability 0.17, control-like (negative SHAP) right pars orbitalis theta in 72% of them (versus 84% of controls and 81% positive in detected tinnitus), projecting to the control side of a control-versus-tinnitus discriminant axis (mean −0.47, versus +1.66 and −1.41). They differ from detected tinnitus only in modestly better high-frequency hearing (HF-PTA Cohen’s *d* = −0.34, *p*_FDR_ = 0.019), with indistinguishable distress (THI and TFI both *p >* 0.70), and the neural difference survived HF-PTA matching at a 10 dB caliper (*d* = −1.27, *p <* 0.0001), indicating tinnitus subtype rather than hearing level. The model thus captures a signature tied to hearing-loss-driven tinnitus, whereas near-normal-hearing but equally distressing tinnitus appears to reach the percept through a distinct, uncharacterised mechanism, with neural habituation a further possibility. Both readings expose a cross-sectional limitation, as deviation is measured once and compensatory reorganisation would attenuate it over time. Psychoacoustics were similarly uninformative (Supplementary Fig. S4): loudness was uncorrelated with predicted probability (*ρ* = −0.009, *p* = 0.897) and SHAP (*ρ* = −0.023, *p* = 0.735), and pitch showed no effect (*H* = 2.83, *p* = 0.243). Resting-state EEG deviation therefore classifies tinnitus at AUC = 0.819 where perceptual reports from the same cohort are inconclusive, and the classifier is ready for proof-of-concept screening evaluation, while still requiring longitudinal designs, richer phenotyping, and an extended marker set.

### F. Preprocessing and multi-site training are decisive

Artefact-rejection stringency showed a non-monotonic, *Goldilocks* relationship with performance: Level 2 (*autoreject* epoch rejection without ICA) outperformed both the minimal Level 1 and the aggressive Level 3. Minimal cleaning left tinnitus power far outside the normative distribution, so the framework acts as an implicit artefact detector, flagging contaminated recordings as extreme deviations rather than silently biasing results, over-aggressive ICA instead discarded genuine theta and alpha signal, costing 0.091 AUC. This reproduces the Level 2 optimum reported by Bomatter et al. [35] across 2600+ recordings, converging on the prescription that moderate automated rejection is optimal while further component removal yields diminishing returns.

Multi-site training is equally decisive. In the 7 × 7 transfer analysis, training on one site and testing at another yielded near-chance AUC (0.45 to 0.66), whereas the same LGBM pipeline reached 0.757 to 0.876 when trained on six sites. The gap reflects insufficient per-site data (median *n ≈* 78) and residual site variance, and an analogous single-to-multi-site gap for EEG classification of consciousness states [16] marks this a general challenge for clinical EEG biomarkers. These results argue for consortium designs as the standard, for which the TIDE protocol provides a template [26]. Harmonisation learned on controls only proved necessary: applied jointly to case-control data, *ComBat* risks absorbing tinnitus-related variance into the site-effect estimate, whereas the controls-only strategy preserves individual deviation while removing technical offsets [17], [21].

### G. Limitations and future directions

Source reconstruction used the *fsaverage* template without individual MRI, introducing localisation error most severe for deep limbic structures where template-to-individual displacement can exceed 10 mm [50], and the beamformer suppresses correlated sources by design, potentially attenuating synchronised bilateral generators. The dominance of *parsorbitalis-rh* warrants caution, as orbitofrontal cortex sits adjacent to the lateral rectus muscle and residual myogenic contamination cannot be excluded without concurrent EMG, future work should couple individual-MRI beamforming to the pipeline and validate top ROIs against MEG. Bayesian linear regression assumes a Gaussian normative distribution, but the positive shift in power Z-scores (median feature mean = 0.711) indicates this is violated under LOSO extrapolation when a site lies outside the training distribution, hierarchical Bayesian regression modelling site as a random effect offers a principled remedy with demonstrated calibration gains [51] and is already present as commented infrastructure in `PCNtoolkit` [52]. With a median of *n*_tin_ *≈* 39 per site, per-site confidence intervals are wide and a single atypical site disproportionately influences the transfer matrix, which a replication cohort from an eighth held-out site would resolve. All analyses are cross-sectional, so the FPN-theta deviation cannot be assigned a causal role, the most consequential open question is whether normative Z-scores track chronification, which longitudinal modelling of Z-score trajectories could test directly, including whether the orbitofrontal theta signature precedes chronification and reverts with intervention. The binary classifier is also agnostic to clinically relevant subtypes (tonal versus noise-like, unilateral versus bilateral, normal-hearing tinnitus, hyperacusis comorbidity), which a one-class normative extension implementable in `PCNtoolkit` could resolve through unsupervised subtyping. Finally, the LOSO AUC of 0.819 approaches but does not meet the threshold for regulatory-grade diagnostic use, which will require the additional site data above and formal validation against pre-specified criteria consistent with FDA/EMA biomarker qualification. We also note that the analysis did not follow a pre-specified statistical analysis plan. Instead, we explored feature representations, connectivity measures, preprocessing levels, and classifiers to identify the model that best differentiated patients from controls. This exploratory strategy introduces analytic flexibility that can inflate apparent performance, classifier folds excluded the test site, but upstream harmonisation and reference-distribution estimation preceded classifier cross-validation, as detailed in SM4. Consequently, these estimates are not fully independent end-to-end validation of the pipeline. A confirmatory study applying the frozen pipeline to a new cohort under a pre-registered analysis plan remains necessary. The approach itself is architecturally transferable to any condition where heterogeneity and multi-site acquisition undermine case-control statistics, as shown in Parkinson’s disease, Alzheimer’s disease [22], and temporal lobe epilepsy [24], and it adds a chronic sensory disorder, dominated by top-down attentional and emotional circuitry rather than a bottom-up auditory signature, to that landscape.

## IV. Materials and methods

### A. Study design and participants

Data were drawn from the TIDE (Tinnitus Detection Project) consortium, a prospective, multicentre case–control study registered at ClinicalTrials.gov (NCT06520865), whose design, inclusion criteria, and biomarker-validation plan are described in detail in the study protocol [26]. The participating sites, in the reporting order used throughout, were the University of Texas at Austin (UTA; Austin, USA), University of Illinois Urbana-Champaign (UIUC; Urbana-Champaign, USA), Trinity College Dublin (TCD; Dublin, Ireland), Brai3n Clinic (BCG; Ghent, Belgium), University Hospital Zurich (UZH; Zurich, Switzerland), University Clinic Tübingen (UKT; Tübingen, Germany), and the Tinnitus Clinic at University Hospital Regensburg (UR; Regensburg, Germany).

Participants were recruited between September 2024 and October 2025. The complete cohort comprised 562 participants; 544 were analysed (268 tinnitus cases and 276 controls; Supplementary Fig. S1). Recruitment, eligibility and exclusions are detailed in Supplementary Methods SM2. Group differences in age, sex and hearing (Table II) were addressed through covariate adjustment.

**TABLE II.** Baseline characteristics of the study population. PTA4, pure-tone average (0.5–4 KHZ), THI, Tinnitus Handicap Inventory, TFI, Tinnitus Functional Index

| Characteristic | Tinnitus (*n* = 268) | Control (*n* = 276) |
|---|---|---|
| Age, yr (mean ± SD) | 49.3 ± 14.7 | 41.6 ± 16.5 |
| Male, n (%) | 141 (52.6%) | 118 (42.8%) |
| Female, n (%) | 127 (47.4%) | 158 (57.2%) |
| PTA4, dB HL (mean ± SD) | 15.3 ± 11.2 | 10.7 ± 8.4 |
| THI (mean ± SD) | 25.8 ± 17.0 | — |
| TFI (mean ± SD) | 30.5 ± 18.6 | — |
| Handedness, % right | 84.5% | 87.1% |

All participants provided written informed consent in accordance with the Declaration of Helsinki [53]; site-specific ethics approvals are listed in Supplementary Methods SM8. No formal a priori power calculation was considered; the protocol power statement was included in error and a correction has been requested (SM5).

*a) Reference standard.:* Cases had chronic, constant, subjective, non-pulsatile primary tinnitus for at least six months. Controls reported no tinnitus lasting longer than two minutes on any single occasion. Status was established by clinical interview; no objective confirmatory test exists. Full reference-standard procedures are given in Supplementary Methods SM2.

### B. EEG acquisition and preprocessing

Resting-state EEG (eyes-open fixation, 5 min) was recorded across seven sites using 64-channel systems (site-specific acquisition hardware and montages are detailed in the supplementary equipment table) and converted to BIDS format [54] prior to preprocessing. Analysts were aware of participant group labels; no analyst blinding was attempted. All data were preprocessed using `MNE-Python` [55]. Site-specific non-EEG channels (physiological sensors, mastoid references, EOG and EMG electrodes) were removed first, and each recording was aligned to its site’s standard electrode layout. UTA recordings, acquired with an EGI geodesic net, were spherically interpolated onto the EasyCap M1 montage to ensure uniform spatial sampling and compatibility with the automated component classifier used in Level 3 (see below) [56]. Three preprocessing levels of increasing stringency were applied to every recording:

1. *Level 1* (lenient baseline): Recordings were resampled to 250 Hz, bandpass-filtered (1– 100 Hz, zero-phase FIR), referenced to the instantaneous average and were segmented into non-overlapping 10-second epochs.
2. *Level 2* (standard, primary analysis): `AutoReject` [34] was applied to Level 1 epochs to identify bad channels and epochs via cross-validated peak-to-peak thresholding. The consensus parameter was optimised over an 11-point grid from 0 to 1, the maximum interpolation count was searched over *{*1, 4, 8*}* channels, and threshold estimation used 5-fold cross-validation. Bad channels were interpolated using spherical splines, epochs that could not be repaired were rejected.
3. *Level 3* (aggressive): Independent component analysis (*FastICA* from *mne.preprocessing.ICA*) was done to Level 2 epochs and components were automatically classified using `MNE-ICLabel` [57].

All non-brain components were removed. Component classifications were visually inspected for each subject to verify that genuinely neural components were not discarded and that artefact components were correctly identified. At UTA, the EGI HydroCel net introduces a residual geodesic reference artefact that *ICLabel* does not reliably flag, accordingly, the first independent component, which consistently captured this artefact across UTA subjects, was additionally excluded regardless of its ICA label.

The primary analysis throughout this paper uses Level 2 epochs. Classification performance across all three levels is compared in section II-I.

### C. Source reconstruction and parcellation

Source reconstruction was performed using the *fsaverage* template brain [50]. A three-layer boundary element model (scalp, outer skull and inner skull,) were used to compute a forward solution (*mne.bem.make_watershed_bem*). Noise covariance was estimated using an ad-hoc diagonal model, which assumes equal and uncorrelated sensor noise (*mne.make_ad_hoc_cov*). A minimum-norm inverse operator was computed using the dSPM method from `mne.minimum_norm. apply_inverse_epochs`. Since all subjects within a given site share the same amplifier and electrode layout, and are mapped to the same *fsaverage* template, a single inverse operator was computed per site and applied to all subjects at that site, avoiding redundant computation while preserving site-specific channel geometry. Label time courses were extracted for each epoch (*mne.extract_label_time_course*) across all 68 parcels of the Desikan–Killiany (aparc) atlas [58] (34 per hemisphere), excluding the medial wall. Within each parcel, vertex-wise source estimates were averaged after flipping vertex signs to align with the label’s dominant orientation. The resulting label time courses formed the basis for all subsequent feature extraction.

### D. Feature extraction

#### Spectral power

Band-limited spectral power was estimated per epoch using multitaper spectral estimation [59]. Ten frequency bands were defined following established EEG band conventions [60]: delta (1–6 Hz), theta (6.5–8.5 Hz), broad alpha (*α*_0_: 8.5–12.5 Hz), alpha-1 (*α*_1_: 8.5–10.5 Hz), alpha-2 (*α*_2_: 10.5–12.5 Hz), broad beta (*β*_0_: 12.5–30 Hz), beta-1 (*β*_1_: 12.5–18.5 Hz), beta-2 (*β*_2_: 18.5–21 Hz), beta-3 (*β*_3_: 21–30 Hz), and gamma (30–40 Hz). Power spectral density was integrated over each band by averaging across epochs, and log-transforming to approximate normality, yielding a 68 × 10 power feature matrix per subject.

#### Functional connectivity

Phase Lag Index (PLI) [61], Phase Locking Value (PLV) [62], and magnitude-squared coherence (COH) [63] were computed per frequency band using complex *Morlet* wavelet convolution (*spectral_connectivity_time* from, `MNE-Connectivity` [32]). PLI was chosen as the primary connectivity metric because its insensitivity to zero-lag contributions from volume conduction and common references makes it robust to EEG-specific artefacts [61]. For 68 source ROIs, pairwise connectivity produced 68 × 67/2 = 2278 unique pairs per band.

#### Node strength metrics

Two aggregate connectivity metrics were derived per ROI per band.

- *Regional node strength* was defined as the sum of PLI (or PLV/COH) values restricted to anatomically adjacent parcels, those sharing a cortical boundary in the Desikan–Killiany atlas on the *fsaverage* surface, as encoded in a precomputed binary adjacency matrix. This metric captures short-range, anatomically constrained synchrony for each ROI.
- *Global node strength* was defined as the sum of PLI (or PLV/COH) values across all other parcels in the graph, irrespective of anatomical adjacency, capturing each ROI’s total long-range network integration.

Both metrics yielded a 68 × 10 matrix per subject, giving three feature matrices in total: spectral power, regional node strength, and global node strength, each entered into its own separate normative model.

#### Aperiodic parameters

The aperiodic (1/*f*) component of the power spectrum reflects local excitation-inhibition (E:I) balance and has been implicated across neurological and psychiatric conditions [64], it was parameterised using FOOOF [65], yielding the slope (exponent) and offset per ROI per epoch, then averaged.

### E. Multi-site harmonisation

Site-induced batch effects in all feature matrices were removed using *ComBat* [66] as implemented in the `neuroHarmonize` library [17]. Critically, harmonisation parameters were learned exclusively on control subjects to prevent tinnitus-related variance from being absorbed into the site-effect estimates, then applied to all subjects (controls and tinnitus). Candidate biological covariates were age, sex, PTA4 and HF-PTA; hearing definitions, available-ear handling and covariate selection are detailed in Supplementary Methods SM3.

### F. Normative modelling

For each of the three feature modalities (spectral power, regional node strength, global node strength) and each feature column (ROI × band), a separate Bayesian linear regression (BLR) normative model was fitted to the control subsample using the `PCNtoolkit` library [67]. The model comprised: a B-spline basis expansion for nonlinear age modelling (degree = 3, *n*_knots_ = 5), heteroskedastic variance (age-dependent residual spread), fixed effects for sex and the selected hearing covariates, and standardised feature inputs.

Hearing-covariate selection is described in Supplementary Methods SM3.

Two complementary fitting procedures were run. (i) A full model was trained on all controls and applied to tinnitus subjects, producing Z-scores that quantify each tinnitus participant’s deviation from the healthy norm: *Z*^(^*^i^*^)^ = (*y*^(^*^i^*^)^ *− µ*^^(^*^i^*^)^)/*σ*^^(^*^i^*^)^. In the full model, site was included as an additional batch-effect term to absorb any residual inter-site variance not fully removed by *ComBat*. (ii) An unbiased LOSO evaluation was run on controls: for each of seven site-folds, the model was trained on the six remaining sites’ controls and applied to the held-out site’s controls, so every control received exactly one Z-score from a model it never trained on. Site was not included as a batch-effect term in the LOSO models for two reasons: *ComBat* harmonisation had already removed systematic inter-site offsets, and more critically, `PCNtoolkit` cannot estimate a batch-effect parameter for a site that was entirely absent from the training fold, which would cause inference to fail for the held-out site. This yielded a fully held-out control Z-score matrix directly comparable to tinnitus Z-scores. Normative calibration quality was verified using a site-stratified 80/20 split of controls, confirming *N* (0, 1) approximation.

### G. Multi-modal diffusive Mahalanobis deviation features

For each (ROI, frequency band) pair (*r, b*) and each subject *i*, the three modality-specific normative Z-scores were stacked into a three-dimensional deviation vector:

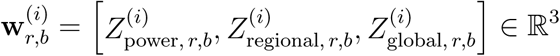

The 3 × 3 covariance matrix **Σ***_r,b_* was estimated from the control subjects’ deviation vectors, with Tikhonov regularisation [68] (**Σ***_r,b_* +10^−4^**I**) to ensure invertibility. The Mahalanobis distance for each subject is then:

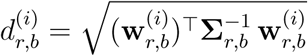

yielding a non-negative scalar that captures the joint deviation across the three neural modalities simultaneously, regardless of the sign of the individual Z-scores. Stacking *d*^(^*^i^*^)^ over all 68 ROIs and 10 frequency bands produced the *band-resolved* feature matrix of 68 × 10 = 680 features per subject, used as the primary input to classification. A band-averaged representation averaging across frequency bands, yielding 68 ROI features, was used for network-level and lateralisation analyses where interpretability required per-ROI rather than per-ROI×band resolution.

### H. Classification

Three classifiers were benchmarked under the same conditions: random forest and radial-basis-function SVM as classical ensemble and kernel-based baselines [69], and LightGBM (LGBM) [31] as a modern gradient-boosted tree framework well-suited to high-dimensional structured data. The primary classifier was selected empirically based on LOSO ablation performance, all three were embedded in a `scikit-learn` [69] pipeline of median imputation (*SimpleImputer*) and Z-score standardisation (*StandardScaler*) with fixed hyperparameters. Cross-validation used *StratifiedGroupKFold* (*k* = 7, grouped by site), ensuring every subject was held out as part of their own site (LOSO). Classification significance was assessed via 100 label-permutation runs under the same splitter, one-tailed *p*-values were computed as the proportion of permuted AUC values exceeding the observed value. An ensemble classifier was constructed by soft-voting (probability averaging) across the top-performing scenarios identified from the ablation.

### I. Ablation scenarios

A systematic scenario comparison was conducted across four axes: (*i*) feature representation (band-averaged 68-dim vs. band-resolved 680-dim), (*ii*) connectivity measure (PLI, PLV, COH), (*iii*) classifier (LGBM, RF, SVM), and (*iv*) preprocessing level (Levels 1–3). Optional hyperparameter optimisation (`Optuna` Bayesian HPO [33], 30 trials per outer fold, 5-fold inner CV) was evaluated for the top scenario. Feature selection variants (*SelectKBest k* = 50, recursive feature elimination, *ElasticNet*) were also compared against the no selection baseline.

### J. Model explainability

*TreeSHAP* values [27] were computed using leave-one-site-out (LOSO) cross-validation (seven folds, one site heldout per fold) to ensure that SHAP attributions reflect genuinely out-of-sample importance. Within each fold, the trained LightGBM model was wrapped in a *TreeExplainer* from the `SHAP` package [30], with interventional feature perturbation and the imputed, standardised training data as background reference. Mean absolute SHAP values were computed across subjects for global feature ranking, additionally, fold-level stability was assessed by computing per-site mean |SHAP| for the top features.

### K. Network and frequency-band analysis

The 68 DK atlas ROIs were mapped to Yeo-7 intrinsic networks [29] (Visual, Somatomotor, Dorsal Attention, Ventral Attention, Frontoparietal, Limbic and Default Mode). Per-subject mean diffusive Mahalanobis scores (band-averaged, 68-feature representation) were aggregated within each network by averaging over member ROIs (both hemispheres). For each of the network × band cells (7 × 10), a Mann-Whitney *U* (*scipy.stats.mannwhitneyu*) test (two-sided) was applied to compare tinnitus and control distributions. *P*-values were corrected for multiple comparisons using the Benjamini-Hochberg false discovery rate (FDR) procedure (*statsmodels.stats.multitest.multipletests*).

### L. Lateralisation analysis

For each of four cortical ROIs spanning the auditory hierarchy and tinnitus-relevant salience circuitry, primary auditory cortex (*transversetemporal*), auditory association cortex (*superiortemporal*), temporoparietal junction (*supramarginal*), and insular cortex (*insula*, included for its role in auditory salience and interoceptive gating rather than as classical auditory cortex, a lateralisation index (LI) was computed per subject:

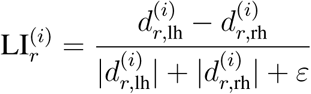

where *ε* = 10^−8^ prevents division by zero and the band-averaged (68-feature) deviation scores are used. LI *>* 0 indicates left-hemisphere dominance, LI < 0 indicates right-hemisphere dominance. Group-level differences in LI were tested with Mann-Whitney *U* (FDR-corrected).

### M. Tinnitus symptom severity regression

To test whether EEG deviation features predicted subjective tinnitus burden, Ridge regression (*α* = 10) and a LightGBM regressor [31] were applied to predict THI and TFI scores from the 680-dimensional deviation features in the tinnitus subgroup (*n* = 268) only. Both models were wrapped in a standardisation pipeline (*StandardScaler* to regressor). Site-stratified cross-validation used *GroupKFold* (grouped by site). The primary metric was the coefficient of determination (*R*^2^) on held-out folds, Spearman *ρ* between individual features and target scores was computed after FDR correction.

### N. Site generalisation

Per-site classification performance was computed from the main LOSO cross-validation predictions, bootstrap 95% confidence intervals for AUC were obtained from 1000 bootstrap resamples of each held-out site’s subjects. A cross-site transfer matrix was constructed by training a random forest classifier on one site and testing on each other site (and itself), producing a 7 × 7 AUC matrix, random forest was used rather than LGBM because per-site training sets (35–90 subjects) are too small for LGBM’s default configuration to generalise reliably. Site pairs with fewer than 10 subjects of either class were excluded.

### O. Statistical analysis

All statistical tests were two-sided unless otherwise noted. FDR correction used the Benjamini-Hochberg procedure (*statsmodels.stats.multitest.multipletests*). Effect sizes for Mann-Whitney tests are reported as rank-biserial correlation. Calibration quality of normative models was assessed by the proportion of control LOSO Z-scores exceeding |1.96| (expected 5.0% under *N* (0, 1)) and by the global mean and standard deviation of control Z-scores.

## Conclusion

We demonstrated that normative modelling of resting-state EEG can detect chronic tinnitus with clinically meaningful accuracy (AUC = 0.832) while remaining interpretable and site-robust. The converging evidence from classification, network analysis, and individual-level deviation mapping points to a coherent picture: chronic tinnitus is dominantly characterised by frontoparietal theta hyperconnectivity anchored in right parsorbitalis. The orthogonality between neural deviation and symptom burden clarifies rather than limits the clinical contribution: we delineate here a reproducible neural phenotype of tinnitus presence that can objectively confirm an internally heard pitched sound independent of subjective report, which is precisely the function of a diagnostic biomarker and a distinct problem from quantifying the burden it produces. Our findings offer a replicable normative framework and a refined pathophysiological target i.e. a orbitofrontal–frontoparietal circuit, for the next generation of tinnitus biomarker development and neuromodulatory intervention.

## Data availability

Anonymized individual participant data (questionnaire, audiometry, EEG) may be made available on reasonable request to the corresponding author, subject to institutional approval and a data sharing agreement. A consortium-wide release is planned once the combined machine-learning analysis of all TIDE paradigms is complete; no date is yet fixed.

## Code availability

All analysis code is openly available at https://github.com/TIDEConsortium/Tinnorm.

## Author contributions

Conceptualization: P.S.S., T.K., F.T.H., J.C., D.D.R., B.L., W.S., S.V., C.R.C., P.K.A.N. Data Curation: P.S.S., M.E., N.W. Formal Analysis: P.S.S. Funding Acquisition: T.K., F.T.H., J.C., D.D.R., B.L., W.S., S.V., C.R.C., P.K.A.N. Investigation: P.S.S., S.A., E.A., F.C., M.E., J.F., N.J., E.K., H.L., L.R.R., E.C.R., N.S., S.M.W., A.Y.-M. Methodology: P.S.S., P.K.A.N. Project Administration: T.K., S.S., F.T.H., J.C., D.D.R., B.L., W.S., S.V., C.R.C., P.K.A.N. Resources: T.K., F.T.H., J.C., D.D.R., B.L., S.V., C.R.C., P.K.A.N. Software: P.S.S., P.K.A.N. Supervision: T.K., F.T.H., J.C., D.D.R., B.L., S.V., C.R.C., P.K.A.N. Validation: M.E., S.S., N.W. Visualization: P.S.S. Writing – Original Draft: P.S.S., P.K.A.N. Writing – Review & Editing: P.S.S., S.A., E.A., F.C., M.E., J.F., N.J., E.K., T.K., H.L., L.R.R., E.C.R., N.S., S.S., S.M.W., A.Y.-M., B.T., N.W., F.T.H., J.C., D.D.R., B.L., W.S., S.V., C.R.C., P.K.A.N.

All authors played a role in writing the manuscript and approved the final version.

