## Supplementary information for "Frontoparietal Hyperconnectivity as a Single-Subject Neural Signature of Chronic Tinnitus"

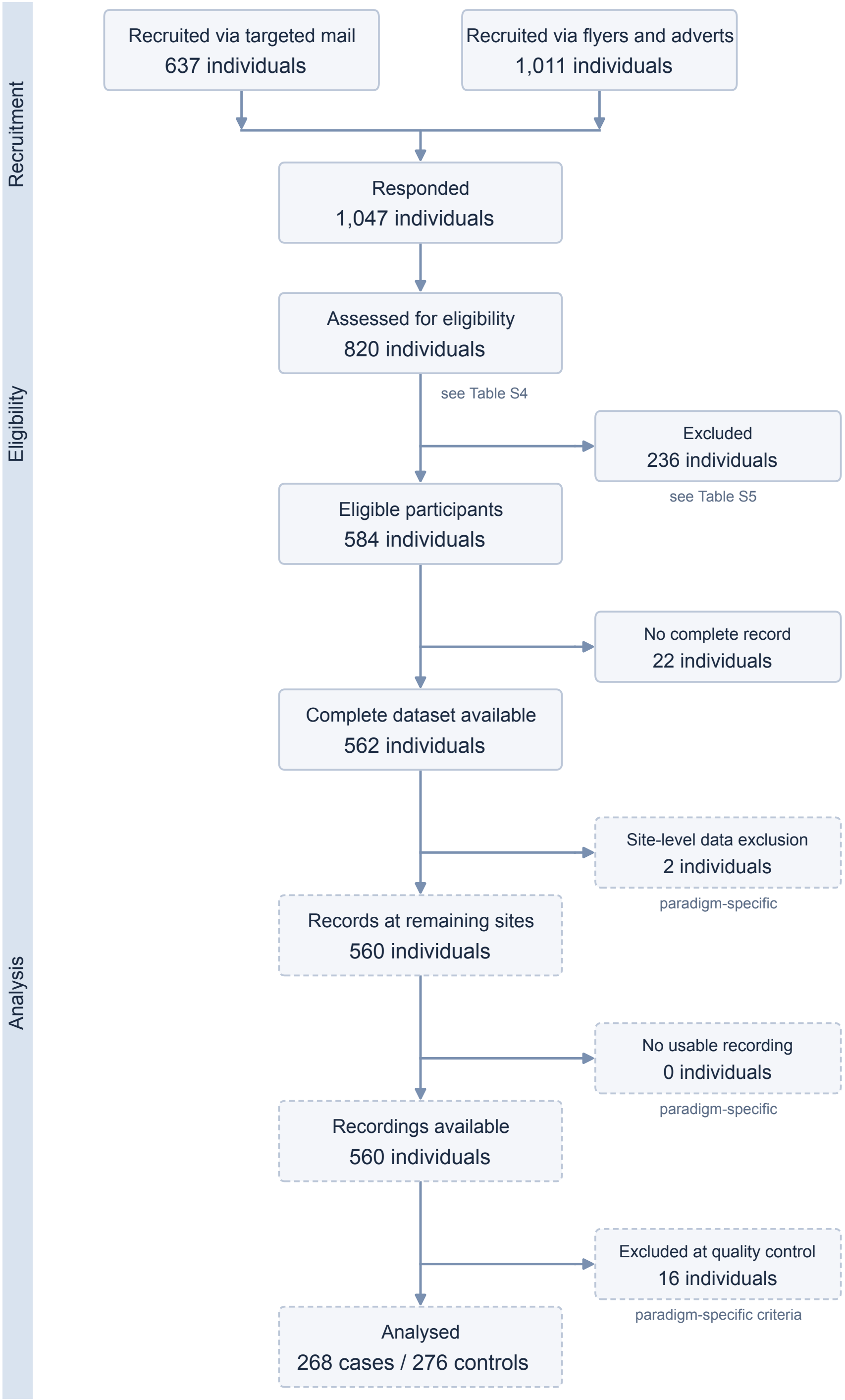


Fig. S1. Participant flow from recruitment to resting-state EEG analysis. The response count was not recorded at UZH, where 117 individuals were assessed for eligibility.

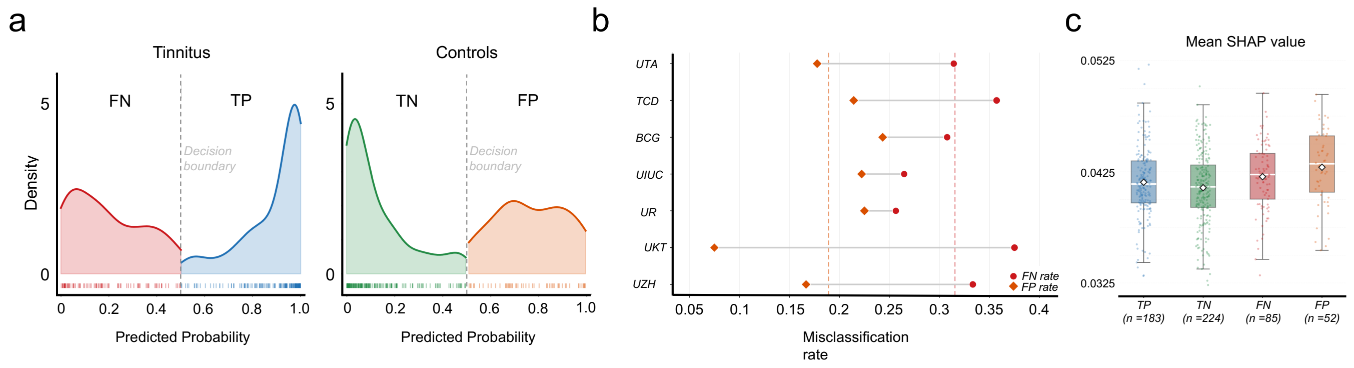


Fig. S2. **Misclassification anatomy of the tinnitus classifier.** (**a**) Kernel density estimate (KDE) of classifier predicted probabilities for each outcome group: true positives (TP, blue) and false negatives (FN, red) within the tinnitus sample; true negatives (TN, green) and false positives (FP, orange) within the control sample. Rug plots mark individual subjects along the horizontal axis; the vertical dashed line denotes the 0.5 decision threshold. Both error groups span the full probability range rather than clustering near the boundary, indicating that a substantial fraction of errors reflect confident mis-assignments rather than marginal uncertainty. (**b**) Per-site false-negative (FN) and false-positive (FP) rates under leave-one-site-out cross-validation. Each site is represented by a dumbbell connecting its FN rate (open circle) and FP rate (filled diamond). UKT shows the largest FN/FP asymmetry, with a markedly low false-positive rate consistent with its overall specificity of 0.925. (**c**) Mean SHAP attribution distributions for all four outcome groups (TP, TN, FN, FP). Boxes span the interquartile range with median bars; individual subjects are overlaid as strip plots. FN subjects exhibit SHAP distributions that overlap with TN rather than TP, indicating that the classifier's internal feature evidence places these tinnitus subjects within the normative brain-deviation range of controls.

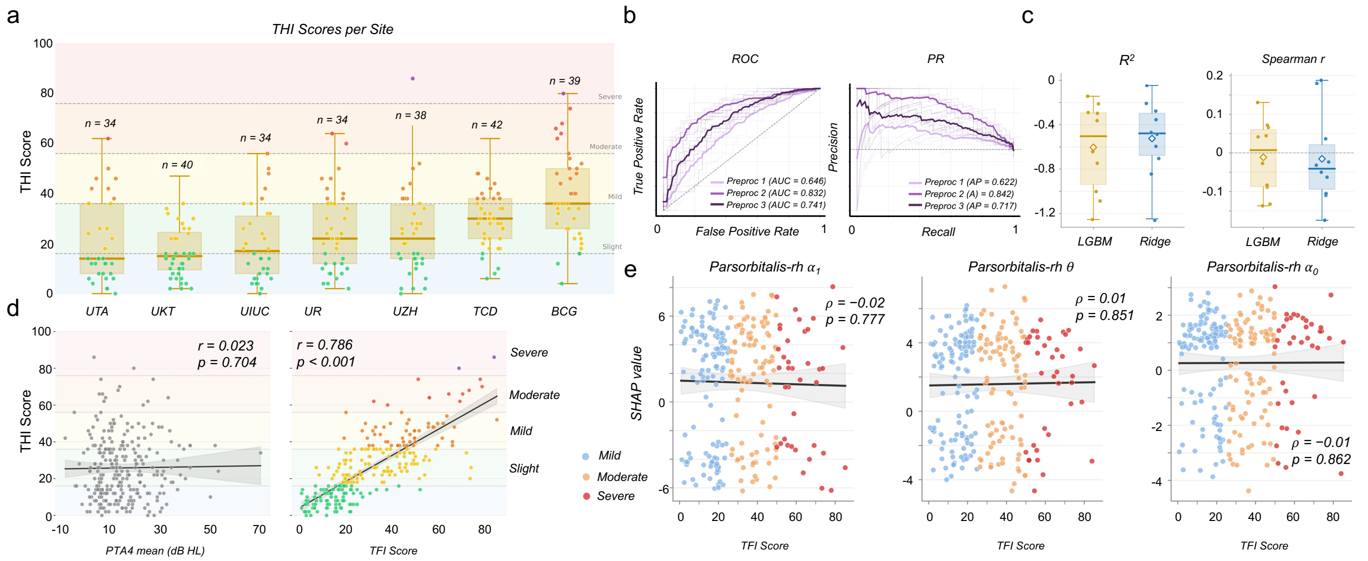


Fig. S3. **Clinical and classification supporting analyses. a**. Per-site THI score distribution for tinnitus participants, sorted by site median. Each column shows a custom box (IQR with median bar and whiskers) overlaid with individual data points. Background bands indicate THI severity categories (slight, mild, moderate, severe, very severe) and dashed horizontal lines mark category thresholds. Sample sizes are annotated above each site. (**b**) Receiver operating characteristic (ROC, left) and precision–recall (PR, right) curves for the three preprocessing levels (1–3) evaluated under leave-one-site-out cross-validation. Faint lines show individual fold curves; bold lines show the cross-fold mean. Dashed reference lines indicate chance performance (AUC = 0.5; baseline precision = 0.5). (**c**) Cross-validation regression performance ($R^2$ and Spearman ρ) for Ridge and LightGBM (LGBM) models predicting THI severity from EEG features. Each strip shows per-fold values from site-grouped cross-validation; boxes span the IQR with median bar; diamonds indicate the cross-fold mean. The dashed line marks zero (no explanatory power). (**d**) Left: scatter of mean four-frequency pure-tone average ($PTA_4$) against THI score in tinnitus participants ($N = 266$; $\rho = 0.02$, $p = 0.70$), with regression line and 95% confidence band. Right: scatter of Tinnitus Functional Index (TFI) against THI in tinnitus participants with available TFI data ($N = 256$; $\rho = 0.79$, $p < 0.001$), illustrating strong cross-instrument convergence. Background bands denote THI severity zones. (**e**) Spearman correlation between SHAP attribution values and TFI score across the five highest-ranked EEG features (by mean SHAP), restricted to tinnitus participants. Dots are coloured by TFI severity (blue: mild < 25; orange: moderate 25–50; red: severe > 50). Solid line shows the OLS regression fit; shading denotes the 95% confidence interval.

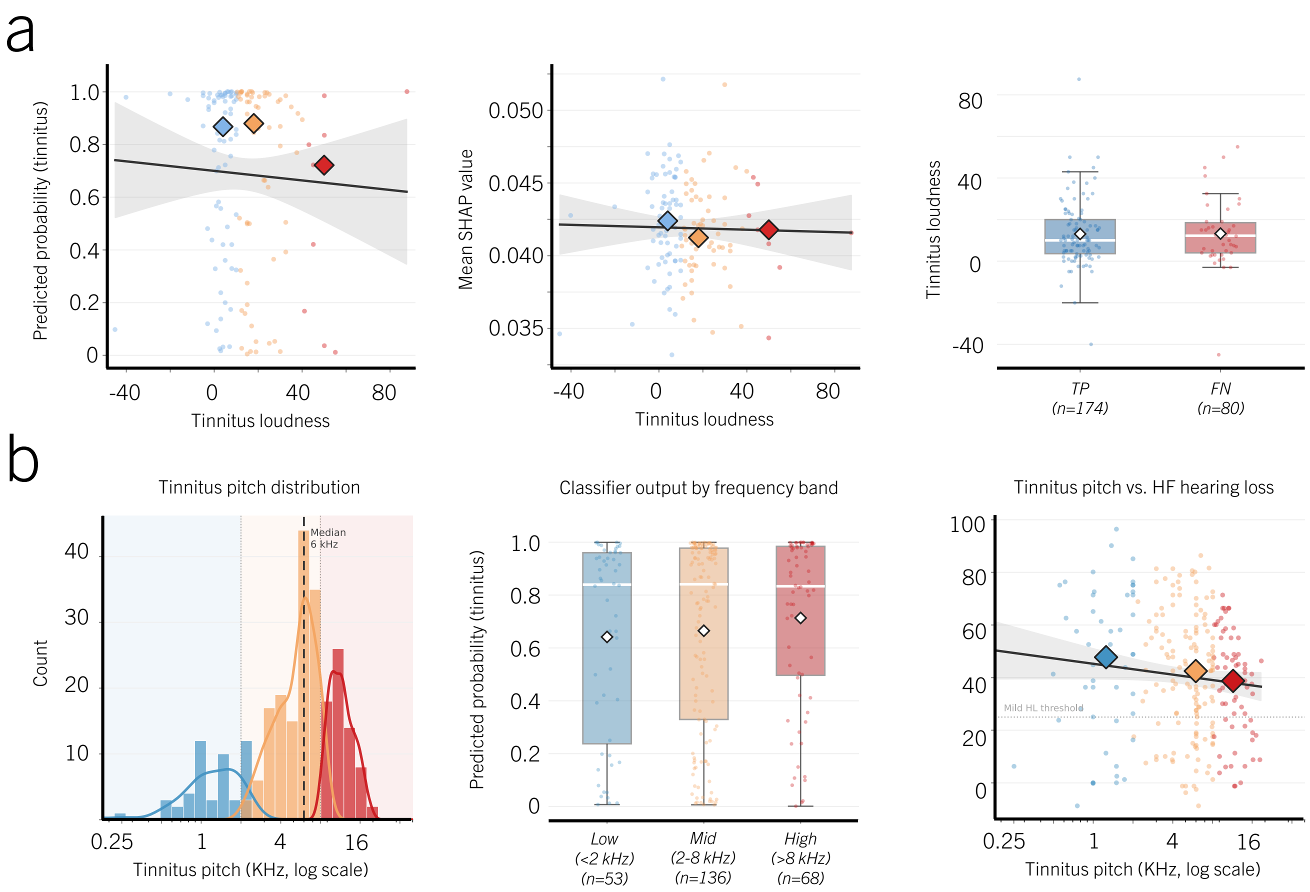


Fig. S4. **Perceptual tinnitus characteristics are not encoded in the EEG classifier.** (**a**) Relationship between audiometrically measured tinnitus loudness (dB SL, mean of right and left ears) and classifier output. *Left*: scatter of LOSO predicted probability versus loudness (Spearman $\rho = -0.063$, $p = 0.460$, n.s.; $n = 140$, loudness in dB SL available for UTA, UR, UIUC and UKT only); *centre*: mean SHAP value versus loudness bin, showing no monotonic trend; *right*: loudness distribution for true-positive (TP) versus false-negative (FN) tinnitus subjects—no group difference was detected (Mann–Whitney $p = 0.801$). These analyses did not establish an association between loudness and the reported classifier outputs. (**b**) Relationship between audiometrically matched tinnitus pitch frequency and classifier output. *Left*: histogram of pitch frequency (kHz, mean of right and left ears where available) with KDE overlays for TP (blue) and FN (red) subgroups; median pitch $\approx$6 kHz. *Centre*: classifier predicted probability across pitch frequency bands (Kruskal–Wallis $H = 1.96$, $p = 0.375$, n.s.). *Right*: scatter of tinnitus pitch frequency versus high-frequency pure-tone average (HF-PTA) with per-band median diamonds; the negative slope ($\rho = -0.152$, $p = 0.014$) may reflect an audiometric matching artefact in which subjects with the greatest high-frequency hearing loss cannot reliably pitch-match to frequencies above their cochlear threshold ceiling, pulling pitch estimates artefactually downward.

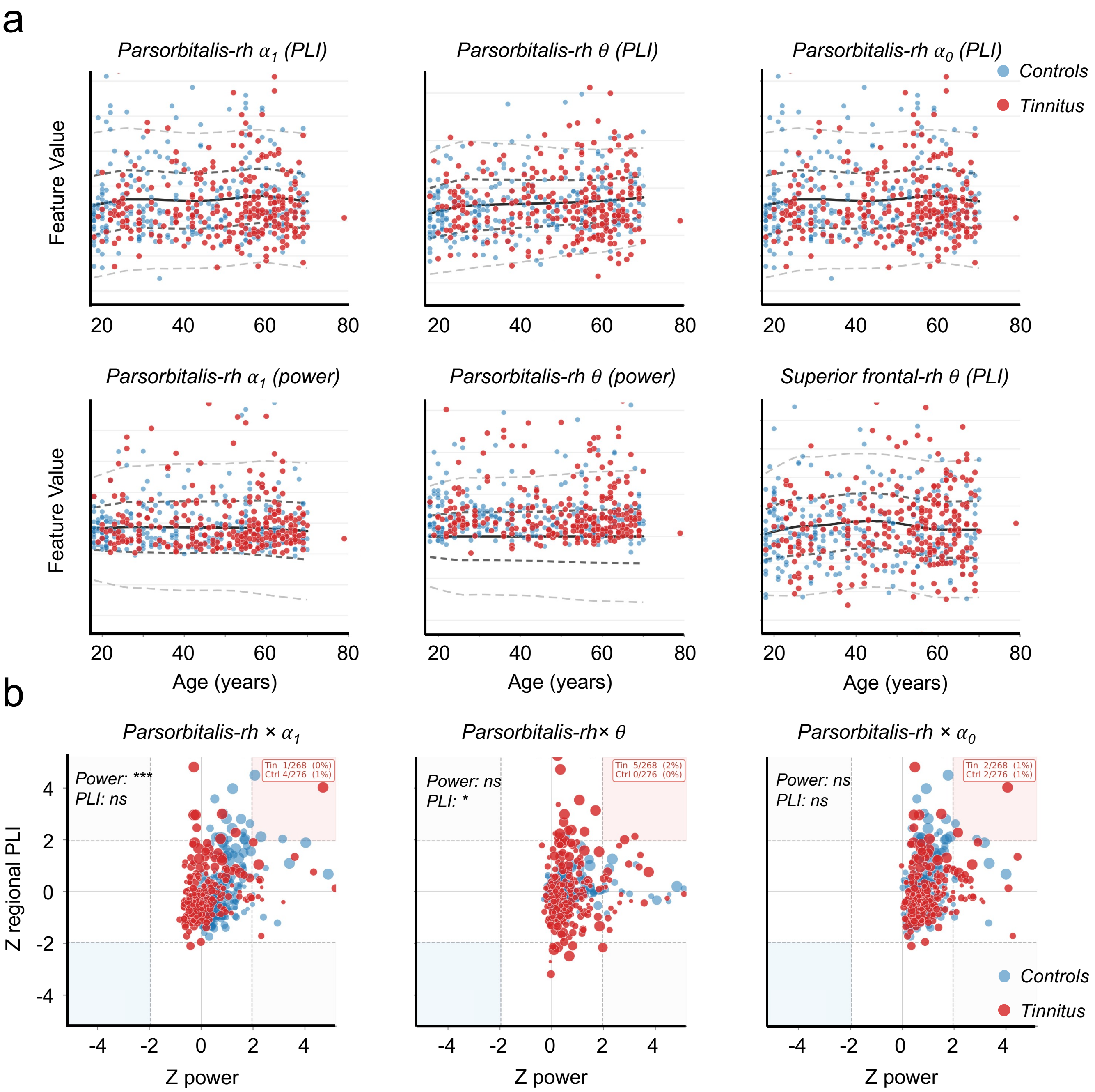


Fig. S5. **Clinical and classification supporting analyses. a**. Normative centile trajectories for the six top-ranked SHAP features across the adult lifespan. Raw harmonised feature values are plotted for controls (blue) and tinnitus participants (red) against age. Smoothed normative percentile boundaries (LOWESS, frac = 0.4) are shown as dashed lines: outer bounds (5th/95th percentile, lighter) and inner bounds (25th/75th percentile, darker); the solid dark line marks the normative median (50th percentile). Each panel corresponds to one ROI × frequency-band × modality combination. (**b**) Two-dimensional Z-score scatter of power versus regional phase-lag index (PLI) deviation for the three top-ranked features (parsorbitalis-RH × $\alpha_1$, θ, and $\alpha_0$). Point size is proportional to the absolute global PLI Z-score (third modality). Controls are shown in blue, tinnitus participants in red. Quadrant shading highlights co-occurring hyperdeviation (both axes above threshold; red zone) and hypo-deviation (blue zone). Dashed lines mark the Z = ±1.96 threshold. Annotated counts show the proportion of each group in the hyperdeviation quadrant.

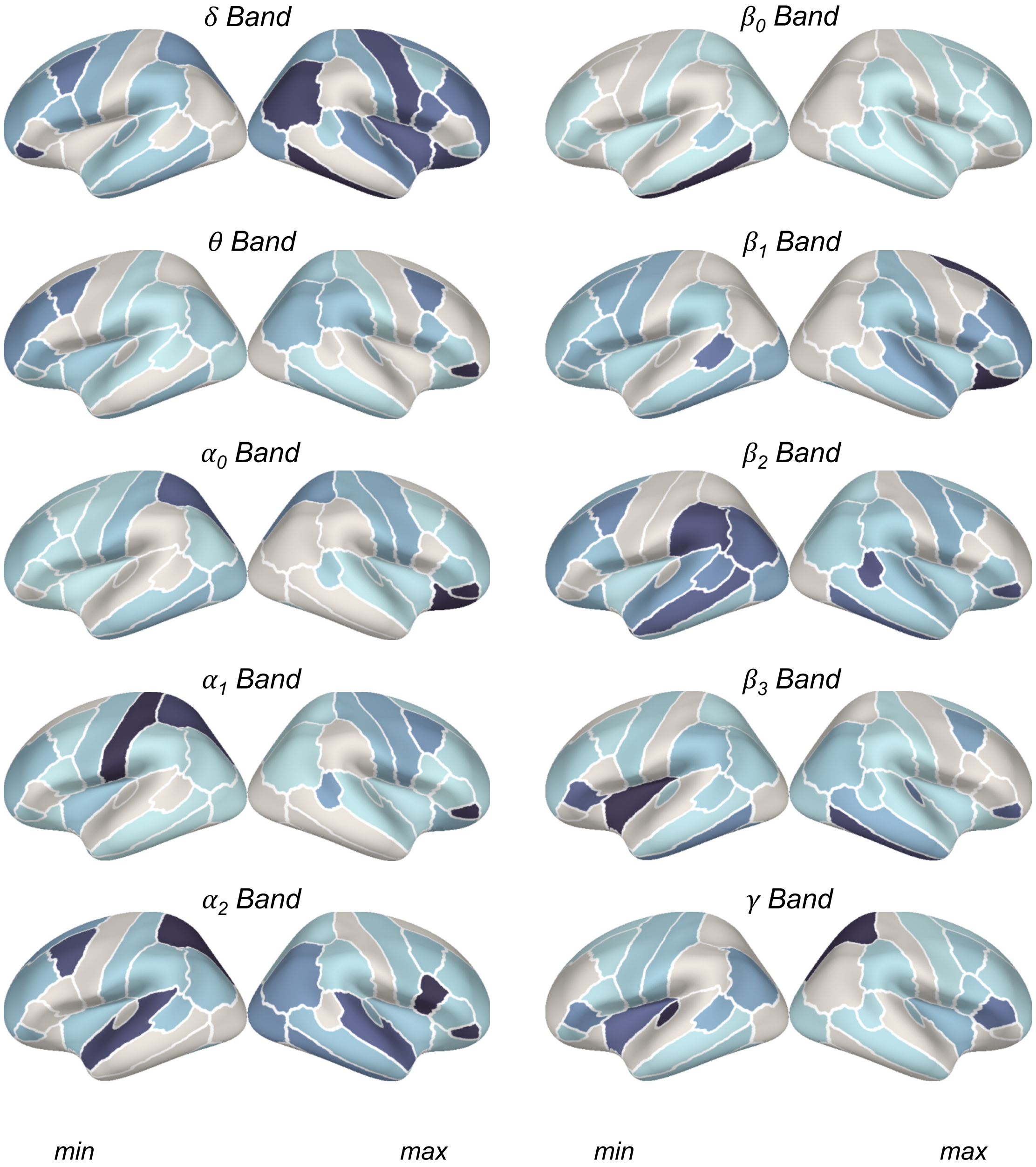


Fig. S6. **Band-resolved SHAP importance projected onto the cortical surface.** Mean |SHAP| per ROI averaged within each frequency band and projected onto the fsaverage inflated surface (DK parcellation). Colour encodes mean |SHAP| normalised per band across ROIs. The *parsorbitalis* (right orbitofrontal cortex) outlier, whose raw mean |SHAP| exceeds all other regions by an order of magnitude in the $\theta$ and $\alpha_1$ bands, is clipped at the 98th percentile to preserve spatial contrast across the remainder of the cortex. In the $\theta$ and lower-alpha ($\alpha_0$, $\alpha_1$) bands, attribution is strongly right-lateralised and confined to prefrontal and orbitofrontal cortex, consistent with the FPN$\times\theta$ signature identified in the main analysis. Upper-alpha ($\alpha_2$) and low-beta bands show a more distributed pattern extending into parietal and temporal association cortex. Delta and gamma carry the lowest total SHAP mass and show no spatially focal pattern, consistent with their absence among the top-ranked features.

### SUPPLEMENTARY METHODS AND REPORTING INFORMATION

#### *SM1. Scope, registration and exploratory status*

This report presents exploratory original research on resting-state EEG features associated with chronic tinnitus. The scientific aims are to investigate individual normative deviations, assess discrimination between the recruited tinnitus and control groups, and characterise the spatial and frequency-specific features contributing to that discrimination. It is not a prospective clinical implementation study, a regulatory biomarker-qualification study, or an independent validation of a previously locked diagnostic product.

The parent TIDE study is a prospective, multicentre case–control study registered at ClinicalTrials.gov (https://clinicaltrials.gov/study/NCT06520865). Prospective refers to participant recruitment and data collection, not to prespecification of the present analytical pipeline. The resting-state paradigm and the machine-learning analyses were not pre-specified in the study protocol or the trial registration and are reported as exploratory. The TIDE protocol is described by Vanneste et al., *BMJ Open* 16:e112788 (2026); it supplies the parent-study context rather than a prespecified analysis plan for the present work.

STROBE is used for the observational study reporting and TRIPOD+AI for the prediction-model development component. A complementary STARD-AI crosswalk is provided for the reported case–control classification results. These reporting frameworks are not interpreted as requiring a different scientific objective or a clinical deployment study. Item-level crosswalks are given in Supplementary Appendices A–C. Applicable reporting gaps are identified explicitly rather than treated as evidence of clinical validation.

The intended current users are researchers with EEG-processing and statistical-modelling expertise. The model outputs are research classification scores; this study does not establish that they are calibrated probabilities of tinnitus in an unselected clinical population. Clinical screening, confirmation of an individual's report, treatment selection and monitoring of treatment response are not validated applications of this work. No prognostic time horizon is defined because the target is contemporaneous group membership rather than a future event.

#### *SM2. Participants, reference standard and sample accounting*

Site codes and institutions are defined in the main Methods; the fixed reporting order is UTA, UIUC, TCD, BCG, UZH, UKT, UR. The five European sites were identified on the basis of expertise and track record in tinnitus research and the two US sites were identified by the European investigators, to enable future implementation under NIH guidelines. Participants were enrolled between September 2024 and October 2025. The study dataset comprised 562 participants: 282 with tinnitus and 280 controls. The recruitment target was 80 participants per site (planned total $N = 560$).

Tinnitus status, the target condition, was chronic, constant, subjective, non-pulsatile primary tinnitus. Case and control status was established at each site by clinical interview, conducted by an audiologist at the US sites and by a physician at the European sites. Cases required a diagnosis of chronic tinnitus of at least six months' duration based on history, perceived as constant, subjective and non-pulsatile, and classified as primary tinnitus. Tinnitus characteristics were documented with ESIT-SQ Part B. Case and control status was established at the eligibility interview, before any EEG was recorded and within the same study visit, so index-test results were not available to the assessors of the reference standard and no intervention occurred between the reference standard and the index test. Controls required no experience of tinnitus lasting longer than two minutes on any single occasion. This is the operationalization of the registered criterion, which excludes anyone who has experienced tinnitus other than short-term. No objective confirmatory test for tinnitus exists, so the reference standard is standardized self-report obtained at clinical interview. This is the limitation the TIDE program sets out to address, and it bounds the accuracy attainable by any index test, since misclassification at the reference standard propagates directly into estimated sensitivity, specificity and discrimination.

Exclusion criteria included objective or primary heartbeat-synchronous tinnitus, otosclerosis, acoustic neuroma, relevant fluctuating ear disorders, acute ear/nose/throat infections, Ménière's disease, vestibular migraine, serious internal, neurological or psychiatric conditions, epilepsy or other central nervous system disorders, clinically relevant drug, medication or alcohol misuse within 12 weeks, severe hearing loss, and single-sided deafness.

Eligibility required age 18–70 years, MoCA $\geq 26$, HQ $\leq 27$, capacity to provide informed consent, and fulfilment of the parent protocol's otological and psychiatric criteria. UZH administered a German version of the HQ questionnaire with three response options per item instead of the four-option version used at the other six sites.

Recruitment sought comparable distributions of age, sex and hearing loss within sites; the achieved groups were not balanced on these variables. Age, sex and hearing measures were therefore included as normative-model covariates. This was not a successfully balanced, individually matched case–control sample. Descriptive characteristics in Table II of the main text should be interpreted as sample descriptions, not as evidence that residual confounding has been eliminated.

*a) Participant accounting.:* The complete cohort comprised 562 participants (282 tinnitus cases and 280 controls). Two participants were excluded post hoc for a combined HADS anxiety and depression score $\geq 14$; no recordings were excluded as unusable, and 16 further participants were excluded at analysis quality control. The final analysis included 544 participants (268 tinnitus cases and 276 controls; Fig. S1).

TABLE S1. Participants in the study dataset by site and group, before analysis-specific filtering.

| Site | Controls | Tinnitus | Total |
|---|---|---|---|
| UTA | 45 | 37 | 82 |
| UIUC | 40 | 43 | 83 |
| TCD | 42 | 41 | 83 |
| BCG | 38 | 39 | 77 |
| UZH | 36 | 42 | 78 |
| UKT | 39 | 40 | 79 |
| UR | 40 | 40 | 80 |
| Total | 280 | 282 | 562 |

*b) Participant selection.:* Questionnaire and audiometry records were matched by participant identifier. Records with missing or invalid required metadata were excluded, together with two participants excluded post hoc for a combined HADS anxiety and depression score $\geq 14$ after passing eligibility screening. This additional criterion was not registered and constitutes a protocol deviation. Subsequent inclusion required available Level-1 sensor-space power features and a non-missing bilateral HF-PTA. These participant-level filters were separate from channel interpolation and epoch rejection during EEG preprocessing.

UTA was excluded from the companion event-related analyses because low evoked-response amplitude and a trigger fault prevented reliable averaging; neither affects resting-state recordings, which require no event markers. This explains the paradigm-specific site inclusion; it does not imply that all UTA recordings were exempt from resting-state quality control.

##### *SM3. Recording and derived EEG variables*

Resting-state EEG was recorded during five minutes of eyes-open fixation, converted to BIDS and processed with MNE-Python. UTA recordings used an EGI HydroCel net and were interpolated to the common EasyCap M1 layout. Site-specific EEG systems, electrode types, cap models, acquisition references, operating systems, PsychoPy versions and sound-calibration equipment are reported in Table S6.

The principal preprocessing level used resampling to 250 Hz, 1–100 Hz zero-phase FIR filtering, average reference, non-overlapping 10-second epochs, and AutoReject epoch rejection/channel interpolation. The comparison levels omitted automated rejection or additionally applied ICA/ICLabel; the primary Level 2 pipeline did not include ICA. Source reconstruction used minimum-norm dSPM on the fsaverage template with an ad-hoc diagonal noise covariance and a site-specific inverse operator. Signals were summarised in the 68 Desikan–Killiany cortical parcels.

For each parcel and frequency band, the three principal modalities were log spectral power, connectivity summed over anatomically adjacent parcels (regional node strength), and connectivity summed over all other parcels (global node strength). PLI, PLV and coherence were compared. Three normative Z-scores were combined using a regularised three-dimensional Mahalanobis distance, yielding 680 parcel-by-band features (68 parcels by ten bands). These are candidate inputs to flexible machine-learning models, not 680 independently estimated logistic-regression coefficients.

TABLE S2. Frequency-band definitions used in the main Methods. Broad bands intentionally overlap their sub-bands; they are not ten disjoint frequency intervals.

| Band | Symbol | Frequency (Hz) |
| --- | --- | --- |
| Delta | $\delta$ | 1–6 |
| Theta | $\theta$ | 6.5–8.5 |
| Broad alpha | $\alpha_0$ | 8.5–12.5 |
| Alpha 1 | $\alpha_1$ | 8.5–10.5 |
| Alpha 2 | $\alpha_2$ | 10.5–12.5 |
| Broad beta | $\beta_0$ | 12.5–30 |
| Beta 1 | $\beta_1$ | 12.5–18.5 |
| Beta 2 | $\beta_2$ | 18.5–21 |
| Beta 3 | $\beta_3$ | 21–30 |
| Gamma | $\gamma$ | 30–40 |

*c) Hearing covariates.:* PTA4 was defined as the mean threshold at 0.5, 1, 2 and 4 kHz; HF-PTA was the mean at 8 and 12.5 kHz at all sites. Ear-specific averages were computed first and then averaged across available ears. When only one ear-specific value was available, that ear was retained; the companion cGPIAS analysis instead excluded participants with one ear missing.

Age, sex, PTA4 and HF-PTA were candidate covariates in both ComBat and normative modelling. If $|r| > 0.85$ between PTA4 and HF-PTA, PTA4 was dropped and HF-PTA retained; otherwise both were used. ComBat evaluated this rule on all merged participants before fitting the harmonisation model on controls. Normative modelling evaluated it on all controls before the 80/20 evaluation split and before the LOSO loop, not separately within each training fold. Missing pairs were omitted from the correlation calculation; this did not itself exclude participants from model fitting.

Participants without an available HF-PTA value were excluded. The remaining hearing summaries were used without additional censoring-specific processing. Where non-responses were coded as the site maximum plus 5 dB, these values were treated as numerical thresholds rather than as censored observations known only to exceed the measurement limit.

*d) Blinding and assessment procedures.:* Analysts were aware of participant group labels; no analyst blinding was attempted. Automated preprocessing was used to limit discretionary processing decisions. No adverse events related to the recordings were reported.

##### SM4. Model development and validation

Random forest, radial-basis-function SVM and LightGBM classifiers were compared across preprocessing levels, connectivity measures and feature representations. The representative single-model configuration and soft-voting ensemble were selected empirically; neither was prespecified. Classifier evaluation used seven site-grouped folds.

*e) Classifier configuration.:* Classification used median imputation, standardisation and, where applicable, feature selection within each classifier-training fold. All three classifiers used balanced class weights and random state 42. Random forest used 300 trees, maximum depth 10, square-root feature sampling and a minimum leaf size of 3. The radial-basis-function SVM used $C = 1$ and $\gamma = 0.001$, with probability estimation enabled. LightGBM used 500 estimators, learning rate 0.05, 31 leaves, unrestricted depth, minimum child size 20, subsampling and feature-sampling fractions of 0.8, and L1/L2 penalties of 0/1.

The principal outer evaluation used `StratifiedGroupKFold`, with recording site as the group and the number of folds equal to the number of sites. Optional Optuna tuning used 30 trials and three-fold stratified inner cross-validation, without site grouping. Soft-voting ensembles averaged scores from the three or five highest-ranked available scenarios, selected using their pooled cross-validation AUC.

Upstream feature construction preceded classifier cross-validation. Missing feature values were replaced by column means before harmonisation. ComBat parameters for the harmonised features were estimated from controls across the available sites and applied to all participants. The alternative residual features were obtained from a separate fit using all participants.

Normative models fitted to all controls generated tinnitus Z-scores, with site included as a batch effect. Control Z-scores were generated by leave-one-site-out normative models without a site batch term. Thus, cases and controls underwent different normative estimation procedures. Before multimodal fusion, Z-scores were clipped to $\pm 10$ and missing Z-scores were replaced by zero. Mahalanobis covariance matrices were estimated from all available controls before classifier evaluation.

Site-grouped cross-validation evaluated classifier performance using precomputed normative features. Harmonisation and reference-distribution estimation were performed before classifier cross-validation; the reported estimates therefore do not constitute fully independent validation of the complete pipeline. Independent replication remains necessary.

##### SM5. Study size and missing-data handling

No formal a priori power calculation was performed. The sample size was determined by the maximum participant enrolment achievable across the seven consortium sites during the recruitment window, and at

544 analysed participants exceeds by an order of magnitude the median cohort of prior single-site EEG tinnitus classification studies. The power statement in the published protocol was included in error and a correction has been requested. The assembled dataset comprised 562 participants: 282 with tinnitus and 280 controls. Classification considered up to 680 candidate features. Dataset totals are not a substitute for analysis-specific denominators after feature availability and filtering. This dimensionality and empirical pipeline selection introduce risks of overfitting and optimistic performance estimates.

Records with missing age, sex, PTA4, site or group were excluded during dataset assembly; subsequent filtering additionally required the initial EEG features and a non-missing HF-PTA. For harmonised power and connectivity features, missing values were replaced by column means before ComBat. During multimodal fusion, missing normative Z-scores were replaced by zero and extreme values were clipped to $\pm 10$. These upstream operations preceded classifier cross-validation. The classifier pipeline additionally included median imputation and standardisation.

Clinical severity analyses were restricted to participants with tinnitus. In the assembled clinical dataset, THI and TFI scores were available for all 282 tinnitus participants. PTA4–THI and TFI–THI clinical pairs were also available for 282 participants (Table S3). These availability counts precede merging with EEG-derived features and do not establish the number included in each plotted or fitted analysis. Regression omitted participants with missing target scores after the clinical–EEG merge; targets were not imputed. TCD loudness ratings were excluded from pooled dB-SL analyses because they used a different measurement scale.

TABLE S3. Clinical-data availability in the assembled tinnitus sample ($n = 282$), before merging with EEG-derived features. PTA4 uses the mean of available ear-specific PTA4 values. These are availability counts, not fitted-analysis denominators.

| Variable or complete pair | Available | Missing |
|---|---|---|
| THI | 282 | 0 |
| TFI | 282 | 0 |
| PTA4–THI | 282 | 0 |
| TFI–THI | 282 | 0 |

*f) Severity-regression configuration.:* Severity regression used band-averaged multimodal deviation features and five-fold cross-validation grouped by site. Ridge regression used $\alpha = 10$; LightGBM regression used 300 estimators, learning rate 0.05, 31 leaves and random state 42. Missing feature values were replaced by zero, and standardisation was fitted within each training fold.

#### *SM6. Classification performance*

Classification performance was summarised using ROC-AUC, sensitivity, specificity, balanced accuracy, and positive and negative predictive values. Threshold-based measures used a classification threshold of

0.5.

Numerical confidence intervals for pooled and ensemble performance are not provided. These estimates should therefore be interpreted as exploratory point estimates, without quantified uncertainty for the complete feature-construction and model-selection procedure. Positive and negative predictive values describe the recruited case–control sample and should not be interpreted as population screening performance.

##### *SM7. Statistical interpretation and assessment not established*

Classification performance was compared with an exploratory reference distribution obtained from 100 label permutations using the same cross-validation splitter. Given the limited permutation count, these comparisons are not used to claim $p < 0.001$ or confirmatory significance for the selected pipeline.

Normative calibration and classification-probability calibration are different assessments. The former evaluates the reference Z-score distribution; a two-sided threshold $|Z| > 1.96$ has nominal probability approximately 5%, while the one-sided threshold $Z > 1.96$ has nominal probability approximately 2.5% under a standard normal distribution. Normative calibration does not demonstrate that a classifier score of 0.8 corresponds to an 80% clinical probability of tinnitus. Classifier calibration is not established by the results reported here. No classifier calibration curve or Brier score is reported; the normative calibration assessment does not replace those evaluations.

Clinical net benefit, comparison of competing patient-management strategies, prospective workflow effects and regulatory qualification are outside the evaluated research questions. No claim about those outcomes is inferred from discrimination. Likewise, variation in performance across sites is not evidence of demographic fairness. Performance equity across sex, age, hearing-loss severity or underrepresented populations has not been established by the reported site analysis; demographic covariate adjustment is not a fairness test.

The absence of successful THI/TFI prediction is evidence that these features did not predict burden under the tested procedures, not proof of statistical independence or biological orthogonality. Non-significant auditory-region findings do not establish normal or intact auditory physiology. Error-defined subgroups and SHAP rankings are exploratory descriptions, not independently established mechanisms or causal effects. SHAP attributes model outputs and cannot rule out residual myogenic contamination or residual confounding. In the reported mean absolute SHAP ranking, right pars orbitalis alpha-1 precedes theta; network-level theta group differences and feature-level model attribution are distinct summaries.

##### *SM8. Reproducibility and research governance*

Analysis code is available at https://github.com/TIDEConsortium/Tinnorm.

*g) Software and reproducibility.:* Analyses were implemented in Python. Classifier configuration, hyperparameter tuning, ensemble selection and cross-validation procedures are described in SM4. Severity-regression configuration and fold-specific standardisation are described in SM5.

The reported results are not linked to a specific repository release or commit. A versioned environment export, a complete run-specific configuration and fitted prediction objects are not provided with this report. The methodological descriptions and available analysis code therefore do not constitute a complete archived record for exact reproduction of the reported predictions. No claim is made that a deployable trained model has been deposited.

*h) Data access.:* Anonymized individual participant data (questionnaire, audiometry, EEG) may be made available on reasonable request to the corresponding author, subject to institutional approval and a data sharing agreement. A consortium-wide release is planned once the combined machine-learning analysis of all TIDE paradigms is complete; no date is yet fixed.

*i) Public involvement.:* Members of the public were not involved in the design, conduct, analysis or reporting of this study.

All participants provided written informed consent in accordance with the Declaration of Helsinki. The study was approved by the responsible ethics committee at each site: UTA (STUDY00004651), UIUC (IRB24-1117), TCD (SPREC022020-07), BCG (6497-B3002024000114), UZH (KEK-ZH 2025-01839), UKT (079/2024BO2), and UR (23-3580-101). Funding, funder involvement and competing interests are reported in the main manuscript.

The pipeline is intended for research use and has not been validated for clinical decision-making. Independent replication and evaluation against non-tinnitus clinical comparison groups are needed to establish generalisability and disorder specificity.

TABLE S4. Recruitment and complete-record availability by site.

| | UTA | UIUC | TCD | BCG | UZH | UKT | UR | All sites |
|---|---|---|---|---|---|---|---|---|
| Recruited via targeted mail | — | — | — | 226 | — | 225 | 186 | 637 |
| Recruited via flyers and adverts | 151 | 336 | 185 | — | 117 | — | 222 | 1,011 |
| Responded | 120 | 302 | 147 | 176 | — | 199 | 103 | 1,047 |
| Assessed for eligibility | 108 | 108 | 130 | 159 | 117 | 95 | 103 | 820 |
| Excluded at eligibility assessment | 25 | 25 | 43 | 76 | 30 | 14 | 23 | 236 |
| Eligible participants | 83 | 83 | 87 | 83 | 87 | 81 | 80 | 584 |
| Without a complete record in the final dataset | 1 | 0 | 4 | 6 | 9 | 2 | 0 | 22 |
| Complete dataset available | 82 | 83 | 83 | 77 | 78 | 79 | 80 | 562 |
| of whom tinnitus cases | 37 | 43 | 41 | 39 | 42 | 40 | 40 | 282 |
| of whom controls | 45 | 40 | 42 | 38 | 36 | 39 | 40 | 280 |

TABLE S5. Eligibility exclusion reasons by site.

| | UTA | UIUC | TCD | BCG | UZH | UKT | UR | All sites |
|---|---|---|---|---|---|---|---|---|
| Tinnitus not chronic | 4 | 4 | — | 4 | 10 | — | — | 22 |
| Age $>$ 70 years | 6 | — | 2 | 5 | 3 | — | 3 | 19 |
| MoCA $<$ 26 | 3 | 1 | — | — | — | 3 | — | 7 |
| HQ $>$ 27 | 3 | 1 | 2 | 2 | — | 5 | 3 | 16 |
| Loudness discomfort level $<$ 70 dB HL[a] | — | 2 | — | — | — | — | — | 2 |
| Objective tinnitus | — | — | — | — | 2 | — | — | 2 |
| Otosclerosis or acoustic neuroma | — | — | — | 3 | — | — | 4 | 7 |
| Ménière's disease | — | — | 2 | 1 | — | — | 1 | 4 |
| Vestibular migraine | — | — | — | 2 | — | — | — | 2 |
| Serious internal, neurological or psychiatric condition | 1 | 1 | — | 6 | 2 | — | 1 | 11 |
| Epilepsy or other disorder of the central nervous system | — | — | 2 | 3 | 1 | — | — | 6 |
| Clinically relevant drug, medication or alcohol misuse | — | — | 1 | — | 1 | — | — | 2 |
| Severe hearing loss | — | 3 | 2 | 5 | 2 | — | 5 | 17 |
| Single-sided deafness | — | — | — | 2 | — | — | — | 2 |
| HADS $>$ 14[b] | 3 | — | — | 1 | — | 3 | 1 | 8 |
| Not matchable to a counterpart | — | 5 | 24 | 18 | — | — | 1 | 48 |
| Lost interest, contact lost or illness | 5 | 8 | 7 | 24 | — | 1 | 4 | 49 |
| Reason not recorded | — | — | — | — | 9 | 2 | — | 11 |
| Additional TCD exclusion (reason unspecified) | — | — | 1 | — | — | — | — | 1 |

[a]Loudness discomfort levels were used only at UIUC as part of routine clinical exclusion. [b]HADS $>$ 14 was initially used for exclusion at several sites and was modified during recruitment. These eligibility criteria are distinct from the two post hoc combined-HADS $\geq$ 14 exclusions reported for the resting-state analysis.

TABLE S6. Site-specific equipment and software across the seven sites, following the shared equipment record. Blank source entries are marked Not recorded.

| Equipment / parameter | UTA | UIUC | TCD | BCG | UZH | UKT | UR |
|---|---|---|---|---|---|---|---|
| Audiometry / Headphones | RadioEar DD450 | HDA200 | HDA300 | HDA200 | HDA200 | HDA300 | HDA300 |
| Audiometry / Audiometer | AudioStar (GSI) | Equinox (Interacoustics) | Hekto (Hörentek) | Not recorded (MedRx) | RME UCX II | AT1000 (Auritec) | Astera 2 (Otometrics) |
| Audiometry / Level step size | 5 dB | 5 dB | 5 dB | 5 dB | 1 dB | 1 dB | 5 dB |
| Audiometry / Max frequency (kHz) and output (dB HL) | 16 (60) | 16 (60) | 12.5 (95) | 16 (60) | 12.5 (90) | 14 (Not recorded) | 16 (40) |
| Audiometry / Tinnitus loudness unit | dB HL and dB SL | dB HL and dB SL | dB HL | dB HL | dB SPL | dB HL | dB HL |
| Audiometry / Tinnitus-matching frequency steps | 50 Hz | 0.25–8 kHz: 1/2 octave; > 8 kHz: 1/6 octave | 25 Hz | 1 Hz | 1 Hz | At PTA frequencies | 10 Hz |
| Audiometry / Tinnitus-matching loudness steps | 1 dB | 2 dB | 5 dB | 1 dB | 1 dB | 1 dB | 5 dB |
| Audiometry / Insert earphones | EARtone GOLD 3A (3M) | EARtone GOLD 3A (3M) | ER2 (Etymotic) | ER2 (Etymotic) | ER2 (Etymotic) | ER2 (Etymotic) | ER2 (Etymotic) |
| Audiometry / Acoustic chamber | Yes | No | No | No | Yes | Yes | No |
| EEG / System | EGI Philips (Magstim) | SynAmps 2 (Neuroscan) | ActiveTwo (BioSemi) | eego mylab (ANT Neuro) | actiCHamp (Brain Products) | actiCHamp (Brain Products) | actiCHamp (Brain Products) |
| EEG / Electrodes | Passive | Passive | Active | Passive | Active | Active | Passive |
| EEG / Cap | HydroCel GSN 64 (EGI) | Quik-Cap Neo Net 64 (Compumedics Neuroscan) | 64-channel (ECI) | waveguard original 64-channel 10/10, shielded (ANT Neuro) | actiCAP slim 64 (Brain Products) | actiCAP slim 64 (Brain Products) | BrainCap TMS 64 (Brain Products) |
| EEG / Reference electrode | Cz | Between Cz and CPz | Cz | CPz | Fz | FCz | FCz |
| EEG / Blink recording | Yes | Yes | No | No | Yes | No | No |
| EEG / Line noise (Hz) | 60 | 60 | 50 | 50 | 50 | 50 | 50 |
| Presentation / OS | Win 10 | Win 10 | Win 10 | Not recorded | Win 11 | Win 10 | Win 10 |
| Presentation / PsychoPy version | 2021.2.3 | 2023.2.3 | 2021.2.3 | 2021.2.3 | 2022.2.5 | 2021.2.3 | 2021.2.3 |
| Presentation / Sound calibration | CAL250 calibrator | B&K 4231 / CAL250 calibrator | GRAS 43AG Ear & Cheek Simulator | GRAS 43AG Ear & Cheek Simulator | B&K 4152 / sound level meter (NTi Audio XL2) | B&K 4152 / KEMAR | Sound level meter (NTi Audio XL2) |
| Presentation / CPU | Intel Core i3-4160 3.60 GHz | Intel Xeon E-2146G 3.50 GHz | Intel Core i5 3.0 GHz | Intel Pentium Gold G5400 3.70 GHz | Intel Core i7 (10th gen) | AMD Ryzen 3 5300U 2.60 GHz | Intel Core i7 2.9 GHz |
| Presentation / RAM | 4 GB | 32 GB | 16 GB | 4 GB | 16 GB | 8 GB | 16 GB |

#### Supplementary Appendix A: STROBE case–control reporting crosswalk

| Item | Reporting item | Location / status | PDF line references |
|---|---|---|---|
| 1a | Study design in title or abstract | Main Abstract | Main: 14–28 |
| 1b | Informative and balanced abstract | Main Abstract | Main: 14–28 |
| 2 | Background and rationale | Main Introduction | Main: 32–73 |
| 3 | Objectives and prespecified hypotheses | Main Introduction; SM1 | Main: 74–87<br>Supp.: 11–21 |
| 4 | Design elements early | Main Methods; SM1 | Main: 419–422<br>Supp.: 16–21 |
| 5 | Settings, locations and dates | Main Methods; SM2 | Main: 422–428<br>Supp.: 34–39 |
| 6a | Eligibility, case ascertainment and control selection | SM2 | Supp.: 40–59<br>Supp.: 69–75 |
| 6b | Matching criteria and controls per case | SM2; group balancing, not individual matching | Supp.: 60–64 |
| 7 | Variables, confounders and diagnostic criteria | Main Methods; SM3 | Main: 486–516<br>Supp.: 40–50<br>Supp.: 97–106 |
| 8 | Data sources and measurement | Main Methods; SM3; Table S6 | Main: 442–484<br>Supp.: 81–100<br>Supp.: Table S6, p. 18 |
| 9 | Measures to address bias | Main Discussion; SM4 | Main: 405–412<br>Supp.: 131–143 |
| 10 | Study-size determination | SM5 | Supp.: 145–153 |
| 11 | Handling of quantitative variables | Main Methods; SM3 | Main: 486–516<br>Supp.: 97–106 |
| 12a | Statistical methods and confounding | Main Methods; SM4 | Main: 561–628<br>Supp.: 101–106<br>Supp.: 120–143 |
| 12b | Subgroups and interactions | Main Results | Main: 171–209 |
| 12c | Missing-data handling | SM5 | Supp.: 154–166 |
| 12d | Handling matching if applicable | Not applicable: no individual matching | Supp.: 60–64 |
| 12e | Sensitivity analyses | Main Results; SM4 | Main: 117–132<br>Main: 211–217<br>Supp.: 116–130 |

Continued on next page

Appendix A (continued)

| Item | Reporting item | Location / status | PDF line references |
|---|---|---|---|
| 13a | Participant numbers at each stage | SM2; Fig. S1; Table S4 | Supp.: 65–75<br>Supp.: Fig. S1, p. 2<br>Supp.: Table S4, p. 16 |
| 13b | Reasons for non-participation | SM2; Table S5 | Supp.: 69–75<br>Supp.: Table S5, p. 17 |
| 13c | Participant-flow diagram | Fig. S1 | Supp.: Fig. S1, p. 2 |
| 14a | Participant characteristics | Main Table II; SM2 | Main: Table II, p. 19<br>Supp.: 60–68 |
| 14b | Missing values for each variable | SM5; Table S3 (partial) | Supp.: 154–166<br>Supp.: Table S3, p. 13 |
| 15 | Exposure summaries | Main Results | Main: 90–114<br>Main: 150–159 |
| 16a | Estimates, precision and adjustment | Main Results; SM6 (pooled precision not reported) | Main: 117–141<br>Supp.: 175–178 |
| 16b | Category boundaries | Main Methods; Table S2 | Main: 486–490<br>Supp.: Table S2, p. 11 |
| 16c | Translation to absolute risk | Not applicable: no relative-risk estimate | — |
| 17 | Other analyses | Main Results | Main: 171–217 |
| 18 | Key results | Main Discussion | Main: 219–227 |
| 19 | Limitations and sources of bias | Main Discussion; SM4; SM7 | Main: 381–412<br>Supp.: 131–143<br>Supp.: 183–202 |
| 20 | Interpretation of results | Main Discussion; SM7 | Main: 405–412<br>Supp.: 196–202 |
| 21 | Generalisability | Main Discussion; SM8 | Main: 393–404<br>Supp.: 225–227 |
| 22 | Funding and funder roles | Main funding statements | Main: 650–658 |

#### SUPPLEMENTARY APPENDIX B: TRIPOD+AI REPORTING CROSSWALK

| Item | Reporting item | Location / status | PDF line references |
|---|---|---|---|
| 1 | Model development/evaluation, population and outcome in title | Main title (partial) | Main: 1–3 |
| 2 | TRIPOD+AI for Abstracts | Main Abstract | Main: 14–28 |
| 3a | Healthcare context and rationale | Main Introduction | Main: 32–73 |
| 3b | Target population, purpose and users | SM1; SM8 | Supp.: 27–32<br>Supp.: 225–227 |
| 3c | Known sociodemographic health inequalities | Not reported | — |
| 4 | Development versus evaluation objectives | Main Introduction; SM1 | Main: 74–87<br>Supp.: 11–21 |
| 5a | Data sources and representativeness | SM1–SM2 | Supp.: 16–21<br>Supp.: 34–39 |
| 5b | Data-collection dates and follow-up | Main Methods; SM2; follow-up not applicable | Main: 428–430<br>Supp.: 37–39 |
| 6a | Study setting and centres | Main Methods; SM2 | Main: 422–428<br>Supp.: 34–39 |
| 6b | Eligibility | SM2 | Supp.: 40–59<br>Supp.: 69–75 |
| 6c | Treatments and their handling | Not reported | — |
| 7 | Preprocessing and quality checking | Main Methods; SM3–SM4 | Main: 442–484<br>Supp.: 81–96<br>Supp.: 131–139 |
| 8a | Outcome definition, rationale and assessment | Main Methods; SM2 | Main: 437–440<br>Supp.: 40–50 |
| 8b | Outcome-assessor qualifications and characteristics | SM2; professions reported | Supp.: 40–44 |
| 8c | Blinding of outcome assessment | SM2; reference standard established before EEG recording | Supp.: 40–50 |
| 9a | Initial predictor selection | Main Methods; SM3 | Main: 486–516<br>Supp.: 91–106 |
| 9b | Predictor definitions and measurement | Main Methods; SM3 | Main: 442–516<br>Supp.: 81–106 |
| 9c | Predictor-assessor qualifications and characteristics | Not reported | — |
| 10 | Study-size determination and adequacy | SM5 | Supp.: 145–153 |

Continued on next page

Appendix B (continued)

| Item | Reporting item | Location / status | PDF line references |
|---|---|---|---|
| 11 | Missing data | SM5 | Supp.: 154–166 |
| 12a | Development/evaluation partitioning | SM4 | Main: 532–544<br>Supp.: 127–143 |
| 12b | Predictor handling and transformations | Main Methods; SM4 | Main: 486–559<br>Supp.: 131–139 |
| 12c | Model building, tuning and internal validation | Main Methods; SM4 | Main: 561–579<br>Supp.: 116–130 |
| 12d | Clustering and heterogeneity | Main Methods; SM4 | Main: 518–544<br>Supp.: 127–143 |
| 12e | Performance measures and comparisons | Main Methods; SM6–SM7; classifier calibration not reported | Main: 616–628<br>Supp.: 172–189 |
| 12f | Model updating | Not applicable: no model updating | — |
| 12g | Predictions from an existing model | Not applicable: no separate existing-model evaluation | — |
| 13 | Class imbalance and recalibration | SM4; recalibration not reported | Supp.: 120–126 |
| 14 | Approaches to model fairness | Not reported; SM7 | Supp.: 190–195 |
| 15 | Model outputs and classification thresholds | SM1; SM6 | Supp.: 27–32<br>Supp.: 172–174 |
| 16 | Development/evaluation data differences | SM2; SM4; partition summaries incomplete | Supp.: 34–39<br>Supp.: 127–143 |
| 17 | Ethical approval and consent | Main Methods; SM8 | Main: 433–434<br>Supp.: 219–224 |
| 18a | Funding and funder role | Main funding statements | Main: 650–658 |
| 18b | Conflicts of interest | Main Competing interests | Main: 660–669 |
| 18c | Protocol access and deviations | SM1–SM2 | Supp.: 16–21<br>Supp.: 65–75 |
| 18d | Registration | Main Methods; SM1 | Main: 419–422<br>Supp.: 16–21 |
| 18e | Data access | Main Data availability; SM8 | Main: 643–646<br>Supp.: 213–216 |
| 18f | Code access and environment | Main Code availability; SM8 | Main: 648<br>Supp.: 204–212 |
| 19 | Patient and public involvement | SM8; public involvement reported | Supp.: 217–218 |
| 20a | Participant flow and outcome counts | SM2; Fig. S1 | Supp.: 65–75<br>Supp.: Fig. S1, p. 2 |

Continued on next page

Appendix B (continued)

| Item | Reporting item | Location / status | PDF line references |
|---|---|---|---|
| 20b | Characteristics and missing data by source/setting | Main Tables I–II; Tables S1/S3 (partial) | Main: Table I, p. 8<br>Main: Table II, p. 19<br>Supp.: Table S1, p. 9<br>Supp.: Table S3, p. 13 |
| 20c | Comparison with development data | Not reported | — |
| 21 | Participants and outcomes in each analysis | Main Methods; SM2; Main Table I (partial) | Main: 428–432<br>Supp.: 65–75<br>Main: Table I, p. 8 |
| 22 | Full model specification and access | SM4; SM8 (partial) | Supp.: 120–139<br>Supp.: 204–212 |
| 23a | Performance and confidence intervals | Main Results; SM6 (pooled precision not reported) | Main: 117–141<br>Supp.: 172–178 |
| 23b | Performance heterogeneity across clusters | Main Results; Main Table I | Main: 134–141<br>Main: Table I, p. 8 |
| 24 | Results of model updating | Not applicable: no model updating | — |
| 25 | Interpretation and fairness | Main Discussion; SM7 | Main: 219–248<br>Supp.: 183–202 |
| 26 | Limitations and generalisability | Main Discussion; SM4; SM8 | Main: 381–412<br>Supp.: 131–143<br>Supp.: 225–227 |
| 27a | Poor-quality or unavailable inputs | SM3; SM5; SM8 (partial) | Supp.: 81–90<br>Supp.: 154–159<br>Supp.: 225–227 |
| 27b | User interaction and expertise | SM1; SM8 | Supp.: 27–32<br>Supp.: 225–227 |
| 27c | Further research | Main Discussion; SM8 | Main: 381–412<br>Supp.: 225–227 |

### Supplementary Appendix C: STARD-AI reporting crosswalk

| Item | Reporting item | Location / status | PDF line references |
|---|---|---|---|
| 1 | AI diagnostic accuracy and an accuracy measure | Main Abstract (partial) | Main: 14–28 |
| 2 | Structured abstract | Main Abstract | Main: 14–28 |
| 3 | Scientific background, intended use and workflow | Main Introduction; SM1 | Main: 32–87<br>Supp.: 27–32 |
| 4 | Objectives and hypotheses | Main Introduction; SM1 | Main: 74–87<br>Supp.: 11–21 |
| 5 | Prospective/retrospective collection | SM1–SM2 | Supp.: 16–21<br>Supp.: 37–39 |
| 6 | Ethics approval | Main Methods; SM8 | Main: 433–434<br>Supp.: 219–224 |
| 7 | Participant-level/data-level eligibility | SM2 | Supp.: 40–59<br>Supp.: 65–75 |
| 8 | Identification of eligible participants | SM2; Table S4 | Supp.: 34–39<br>Supp.: Table S4, p. 16 |
| 9 | Setting, location and dates | Main Methods; SM2 | Main: 422–428<br>Supp.: 34–39 |
| 10 | Consecutive, random or convenience series | Not stated | — |
| 11 | Data source and collection purpose | SM1–SM2 | Supp.: 11–21<br>Supp.: 34–39 |
| 12 | Dataset annotation and annotators | SM2; clinical reference assessment | Supp.: 40–50 |
| 13 | Capture devices and software versions | SM3; Table S6 | Supp.: 81–84<br>Supp.: Table S6, p. 18 |
| 14 | Acquisition and preprocessing | Main Methods; SM3 | Main: 442–484<br>Supp.: 81–96 |
| 15a | Index-test specification | Main Methods; SM4 | Main: 486–579<br>Supp.: 120–139 |
| 15b | Development, training, validation and testing | SM4; partition sample sizes partially reported | Supp.: 116–143 |
| 15c | Index-test thresholds and prespecification | SM1; SM6 | Supp.: 16–21<br>Supp.: 172–174 |
| 15d | Intended users and expertise | SM1; SM8 | Supp.: 27–32<br>Supp.: 225–227 |

Continued on next page

Appendix C (continued)

| Item | Reporting item | Location / status | PDF line references |
|---|---|---|---|
| 16a | Reference-standard procedures | Main Methods; SM2 | Main: 437–440<br>Supp.: 40–50 |
| 16b | Reference-standard rationale | SM2 | Supp.: 47–50 |
| 16c | Reference categories and cut-offs | SM2 | Supp.: 40–46 |
| 17a | Information available to index-test assessors | Main Methods; SM3; analysts unblinded | Main: 444–445<br>Supp.: 111–114 |
| 17b | Information available to reference assessors | SM2; index-test results not available | Supp.: 40–50 |
| 18 | Accuracy estimation and comparison | Main Methods; SM6–SM7 | Main: 616–628<br>Supp.: 172–189 |
| 19 | Handling of indeterminate results | Not explicitly reported | — |
| 20 | Missing index-test/reference data | SM2; SM5 | Supp.: 69–75<br>Supp.: 154–166 |
| 21 | Accuracy variability and prespecification | Main Results; SM4 | Main: 117–148<br>Supp.: 116–130 |
| 22 | Sample-size determination | SM5 | Supp.: 145–153 |
| 23 | Error analysis, bias and fairness | Main Results; Fig. S2; SM7; fairness not reported | Main: 139–141<br>Supp.: Fig. S2, p. 3<br>Supp.: 190–202 |
| 24 | Participant-flow diagram | Fig. S1; SM2 | Supp.: Fig. S1, p. 2<br>Supp.: 65–75 |
| 25 | Development/test-set characteristics | Main Tables I–II; Tables S1/S6 (partial) | Main: Table I, p. 8<br>Main: Table II, p. 19<br>Supp.: Table S1, p. 9<br>Supp.: Table S6, p. 18 |
| 26a | Target-condition severity | Main Table II; Fig. S3 | Main: Table II, p. 19<br>Supp.: Fig. S3, p. 4 |
| 26b | Alternative diagnoses in reference-negative participants | Not reported | — |
| 27 | Test/reference interval and interventions | SM2; same visit, no intervening intervention | Supp.: 40–50 |
| 28 | Target-condition representativeness | SM2; SM6 | Supp.: 34–64<br>Supp.: 175–178 |
| 29 | Differences in external evaluation data | Not applicable: no independent external dataset | — |

Continued on next page

Appendix C (continued)

| Item | Reporting item | Location / status | PDF line references |
|---|---|---|---|
| 30 | Index/reference cross-tabulation | Not reported | — |
| 31 | Accuracy estimates and precision | Main Results; SM6 (pooled precision not reported) | Main: 117–141<br>Supp.: 175–178 |
| 32 | Adverse events | SM3; none reported | Supp.: 113–114 |
| 33 | Limitations, bias and generalisability | Main Discussion; SM4; SM7 | Main: 381–412<br>Supp.: 131–143<br>Supp.: 183–202 |
| 34 | Implications, intended use and clinical role | Main Discussion; SM1; SM8 | Main: 219–248<br>Supp.: 27–32<br>Supp.: 225–227 |
| 35 | Ethical standards, bias and fairness | SM7–SM8 (partial) | Supp.: 190–195<br>Supp.: 219–227 |
| 36 | Registration | Main Methods; SM1 | Main: 419–422<br>Supp.: 16–21 |
| 37 | Protocol access | SM1 | Supp.: 16–21 |
| 38 | Funding and role of funders | Main funding statements | Main: 650–658 |
| 39 | Commercial interests | Main Competing interests | Main: 660–669 |
| 40a | Data/code availability and reuse | Main availability statements; SM8 | Main: 643–648<br>Supp.: 204–216 |
| 40b | Auditable model outputs | SM8; fitted prediction objects not provided | Supp.: 208–212 |
